# Large variation in the association between seasonal antibiotic use and resistance across multiple bacterial species and antibiotic classes

**DOI:** 10.1101/2020.12.21.20248670

**Authors:** Daphne S. Sun, Stephen M. Kissler, Sanjat Kanjilal, Scott W. Olesen, Marc Lipsitch, Yonatan H. Grad

## Abstract

Understanding how antibiotic use drives resistance is crucial for guiding effective strategies to limit the spread of resistance, but the use-resistance relationship across pathogens and antibiotics remains unclear. We applied sinusoidal models to evaluate the seasonal use-resistance relationship across 3 species (*Staphylococcus aureus, Escherichia coli*, and *Klebsiella pneumoniae*) and 5 antibiotic classes (penicillins, macrolides, quinolones, tetracyclines, and nitrofurans) in Boston, Massachusetts. Use of all 5 classes and resistance in 9 of 15 species-antibiotic combinations showed statistically significant amplitudes of seasonality (false discovery rate < 0.05). While seasonal peaks in use varied by class, resistance in all 9 species-antibiotic combinations peaked in the winter and spring. The correlations between seasonal use and resistance thus varied widely, with resistance to all antibiotic classes being most positively correlated with use of the winter-peaking classes (penicillins and macrolides). These findings challenge the simple model of antibiotic use independently selecting for resistance and suggest that stewardship strategies will not be equally effective across all species and antibiotics. Rather, seasonal selection for resistance across multiple antibiotic classes may be dominated by use of the most highly prescribed antibiotic classes, penicillins and macrolides.

## Introduction

Antibiotic resistance is a growing threat to society, with important public health [1] and economic consequences [2]. Antibiotic use is considered a primary driver of resistance not only in the pathogen targeted by the antibiotic but also in host-associated bacteria subject to ‘bystander selection’ [3]. As such, stewardship programs to reduce overall antibiotic prescribing have become a popular strategy for broadly reducing the burden of resistance. However, the efficacy of stewardship efforts has varied widely and, in some cases, shown a limited impact on reducing rates of resistance [4,5]. These findings reflect the complexity of the antibiotic use-resistance relationship, underscoring the need to characterize this relationship across a wide range of bacterial species and antibiotics and identify factors that influence the strength of this association.

Temporal studies have shown that an association between population-level antibiotic use and resistance can be detected on rapid timescales, where seasonal fluctuations in use have been accompanied by seasonal fluctuations in resistance with up to a few months lag [6–9]. To interpret the lag between seasonal use and resistance, Blanquart et al. proposed a model for the relationship between short-term sinusoidal fluctuations in antibiotic use and resistance [10]. This model predicts that antibiotic use determines the rate of change of resistance, such that the derivative of the prevalence of resistance should depend on the level of use. Thus, if antibiotic use follows a sine function over a 12-month period, then peak resistance should lag peak use by a quarter period, or 3 months. The lag can be shortened by including a ‘stabilizing force’ in the model to account for forces that counteract the effect of use and drive fluctuations in resistance towards equilibrium.

Findings from previous seasonality studies have been largely consistent with this model. Studies that focused on antibiotics with wintertime peaks in use (e.g. penicillins, macrolides, and quinolones) have identified positive associations with winter/spring peaks in resistance lagged by 0-3 months in *Streptococcus pneumoniae* [6], *Escherichia coli* [7,8], *Staphylococcus aureus* [7], and *Neisseria gonorrhoeae* [9]. In contrast, a study from the Netherlands that analyzed antibiotics with summer/autumn peaks in use (e.g. Nitrofurantoin, Fosfomycin, Trimethoprim) found that resistance in *E. coli* and *Klebsiella pneumoniae* still peaked in the winter/spring and lagged use by 3-6 months [11], inconsistent with the Blanquart et al. model. The authors of this study attribute the longer lag time to the weaker seasonal fluctuations and lower overall rates of antibiotic use in their study population. However, it is unclear whether resistance to other antibiotics with winter peaks in use would exhibit similarly long lag times in this population, or whether these findings reflect a broader phenomenon where despite different seasonal patterns of use, resistance always peaks in the winter/spring due to other ecological factors.

We aimed to characterize the seasonal relationship between antibiotic use and resistance across antibiotic classes with winter, summer, and biannual peaks in use [6,7,9,12] in Boston, Massachusetts. We studied three clinically relevant species—*Staphylococcus aureus, Escherichia coli*, and *Klebsiella pneumoniae*—which represent skin/nasal and gut colonizing bacteria [13,14] that cause a diversity of infections types and are subject to strong bystander selection [3]. We obtained antibiotic use data from a centralized state-wide insurance claims database and resistance data from two major Boston-area hospitals. Given the near-universal health insurance coverage in Massachusetts [15], this analysis provided a unique opportunity to characterize the antibiotic use-resistance relationship in a dataset that captures nearly all antibiotic use in a population.

## Results

### Seasonality in antibiotic use varies across classes

The five antibiotic classes included in this study each displayed statistically significant seasonal patterns of use (**Fig 1A**). Penicillins and macrolides were most frequently prescribed, with year-round averages of 4.8 and 4.1 daily claims per 10,000 people, respectively. Quinolones, tetracyclines, and nitrofurans were prescribed with year-round averages of 1.8, 1.0, and 0.5 daily claims per 10,000 people, respectively.

**Fig 1.**
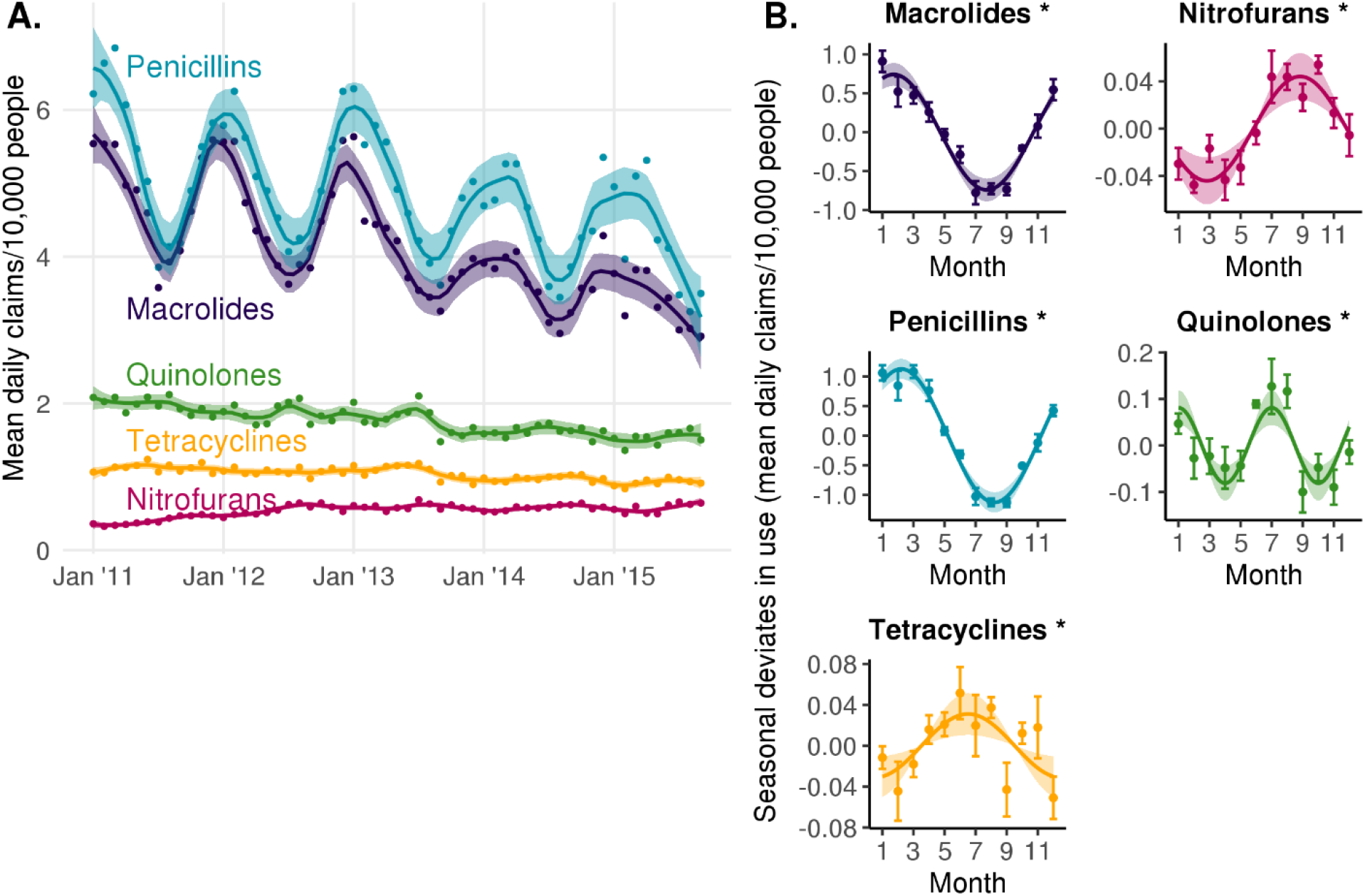
Seasonal patterns of antibiotic use by class. (A) Average daily antibiotic claims per 10,000 people by calendar month in Boston, MA from 2011 to 2015. Lines indicate LOESS smoothing curves and shaded regions indicate 95% confidence intervals. (B) Sinusoidal model fits for monthly prescribing rate. Points indicate monthly mean seasonal deviates in average daily antibiotic claims per 10,000 people by calendar month and error bars indicate the standard error of the mean. Lines indicate the point estimate for the amplitude and phase of the sinusoidal model. Shaded regions indicate the 95% confidence intervals for the amplitude. Asterisks indicate the amplitude of seasonality is statistically significant (FDR < 0.05).

Penicillins had the greatest magnitude change in prescribing rate across seasons, with the seasonal component having an amplitude of 1.1 additional daily claims per 10,000 people (peak to mean) (95% CI, 0.96 to 1.3). This was followed by macrolides (amplitude, 0.74; 95% CI, 0.59 to 0.89), quinolones (amplitude, 0.081; 95% CI, 0.04 to 0.12), nitrofurans (amplitude, 0.04; 95% CI, 0.02 to 0.06), and tetracyclines use (amplitude, 0.03; 95% CI, 0.01 to 0.05) (**Fig 1B**).

The timing of peak prescribing varied by antibiotic class (**Fig 1B**). Macrolide and penicillin use peaked in the winter, around late January (phase, 1.7 months; 95% CI, 1.3 to 2.1; note that phase is indexed to 1.0 representing January 1st) and early February (phase, 2.2 months; 95% CI, 2.0 to 2.5), respectively. Tetracycline and nitrofuran use peaked in the summer, around mid-June (phase, 6.5 months; 95% CI, 4.6 to 8.5) and late August (phase, 8.8 months; 95% CI, 8.2 to 9.4), respectively. Finally, quinolone use peaked twice a year in early January and early July (phases, 1.0 (95% CI, 0.6 to 1.5) and 7.0 (95% CI, 6.6 to 7.5) months).

### Seasonality in antibiotic resistance is prevalent across species and antibiotic classes

Resistance was seasonal for 9 out of 15 species-antibiotic combinations (**Fig 2, S1 Fig, S2 Fig**), with statistically significant amplitudes of seasonality (FDR < 0.05) ranging from a peak log_2_(MIC) increase of 0.028 to 0.063 above the yearly average (**Fig 3**). Ciprofloxacin resistance and nitrofurantoin resistance were seasonal in all three species with a 12-month period (**Fig 2, S1 Fig, S2 Fig**). Resistance to erythromycin in *S. aureus* was also seasonal with a 12-month period (amplitude, 0.048; 95% CI, 0.012 to 0.083). Conversely, tetracycline resistance was not seasonal in any of the three species (**Fig 2, S1 Fig, S2 Fig**). The seasonal patterns of resistance to penicillin class antibiotics were variable across species. Oxacillin resistance in *S. aureus* was seasonal with a 12-month period (amplitude, 0.031; 95% CI, 0.009 to 0.054), while both penicillin resistance in *S. aureus* (amplitude, 0.010; 95% CI, -0.007 to 0.027) and amoxicillin/clavulanate resistance in *E. coli* (amplitude, 0.010; 95% CI -0.0005 to 0.021) and *K. pneumoniae* (amplitude, 0.034; 95% CI, 0.001 to 0.067) did not meet our criterion for seasonality. Ampicillin resistance in *E. coli* was the only species-antibiotic combination with a statistically significant amplitude (0.034; 95% CI, 0.002 to 0.049) that showed a 6-month period in seasonality. However, despite having a slightly worse fit, the 12-month period model of ampicillin resistance in *E. coli* also indicated seasonality (amplitude, 0.041; 95% CI, 0.019 to 0.062) (**S3 Fig**).

**Fig 2.**
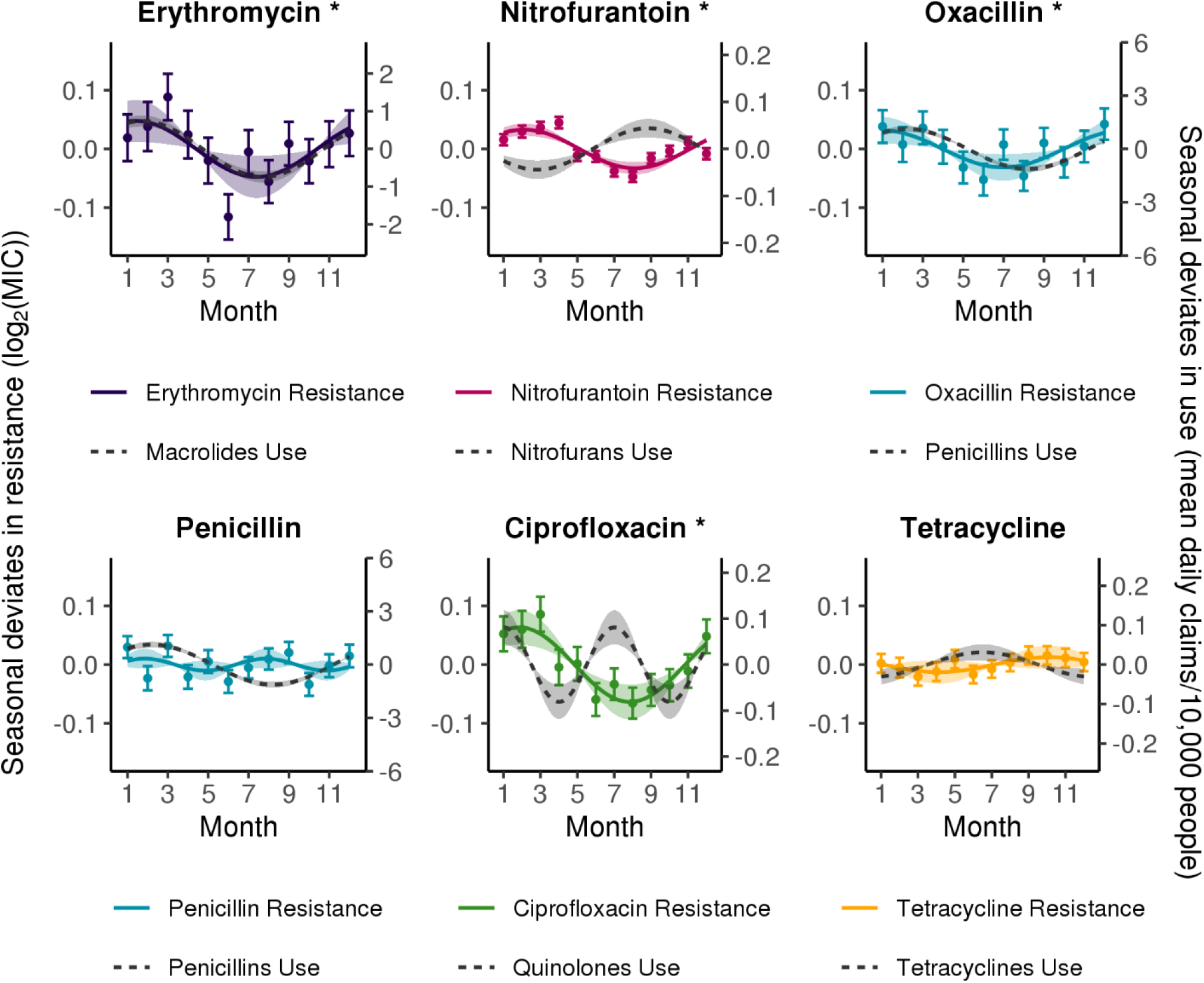
Seasonality of antibiotic use and resistance by class in *Staphylococcus aureus*. Solid lines indicate point estimates of the amplitude and phase from the best-fitting sinusoidal model of resistance (comparing 6- and 12-month periods) to each antibiotic, colored by class. Dashed grey lines indicate point estimates of the amplitude and phase of sinusoidal models for use of the corresponding antibiotic class. Shaded regions indicate the 95% confidence intervals for the amplitude. Points indicate the monthly mean seasonal deviates in resistance and error bars indicate the standard error of the mean. Asterisks indicate the amplitude of seasonality in resistance is statistically significant (FDR < 0.05).

**Fig 3.**
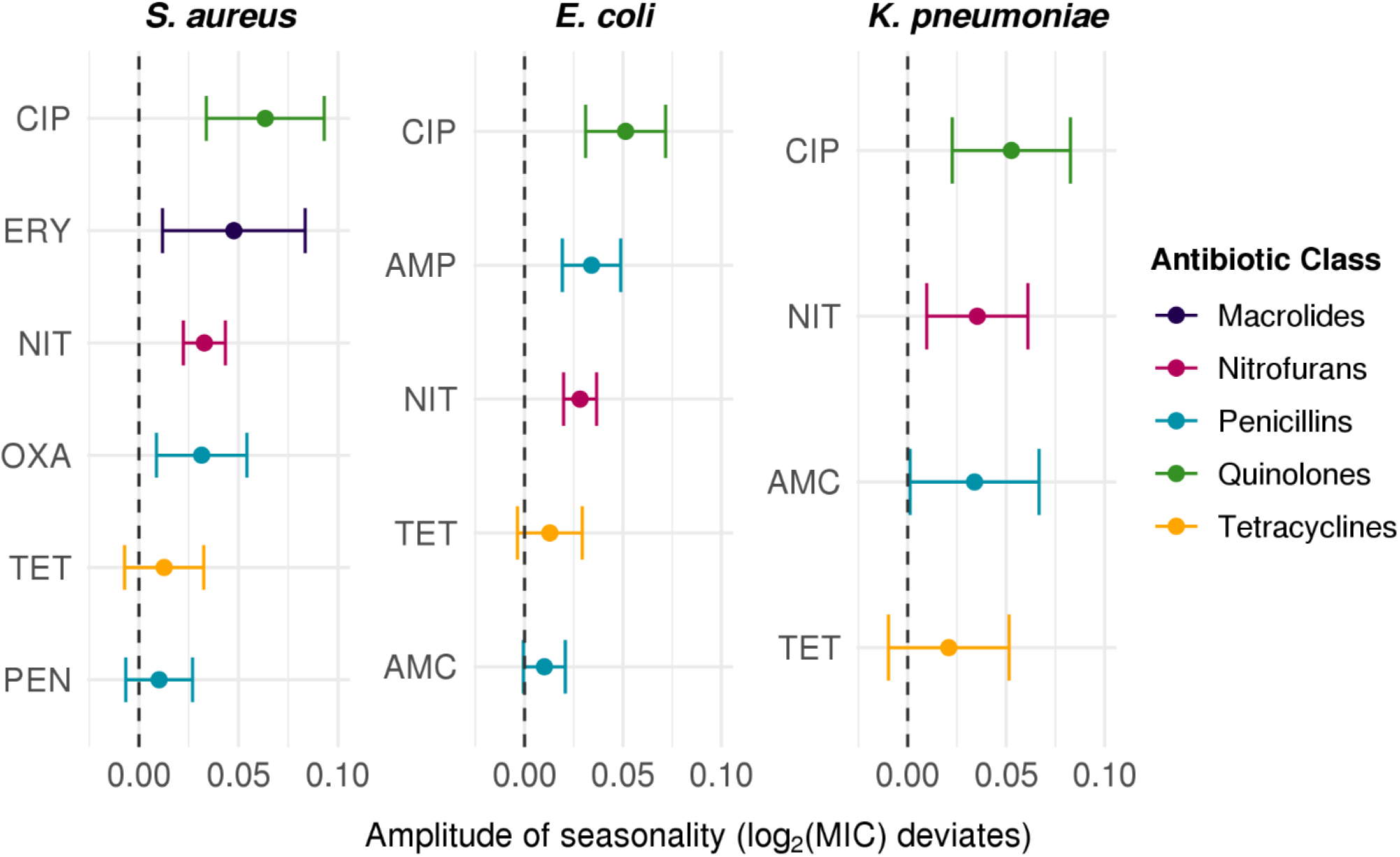
Amplitudes of seasonality of resistance by species and antibiotic class. Comparison of amplitudes estimated from best-fitting sinusoidal models of resistance (comparing 6- and 12-month periods) across antibiotics in *Staphylococcus aureus, Escherichia coli*, and *Klebsiella pneumoniae*. Error bars indicate 95% confidence intervals of the amplitude. Point color indicates the antibiotic class. AMC, Amoxicillin-Clavulanate; AMP, Ampicillin; CIP, Ciprofloxacin; ERY, Erythromycin; NIT, Nitrofurantoin; OXA, Oxacillin; PEN, Penicillin; TET, Tetracycline.

Resistance peaked in the winter to spring months in all 9 seasonal species-antibiotic combinations, with peaks ranging from early December to mid-April (**Fig 4**). Comparing across species, resistance in *E. coli* (median phase, 3.2; range, 2.5 to 4.5) tended to peak slightly later in the year than resistance in *S. aureus* (median phase, 1.6; range, 0.98 to 2.1) and *K. pneumoniae* (median phase, 1.1; range, 0.0 to 2.2). Peak resistance to macrolides and penicillins in *S. aureus* and the first peak in resistance to ampicillin in *E. coli* occurred around the same time of year as peak use of macrolides and penicillins, with lags of -1.2 to 2.3 months. However, resistance to ampicillin in *E. coli* also peaked a second time during the year in October, though use did not. Resistance to nitrofurans in all 3 species peaked between 3.2 to 5.7 months after peak nitrofuran use. Finally, resistance to quinolones in all 3 species peaked once a year about 0.8 to 2.2 months after the first peak in quinolone use.

**Fig 4.**
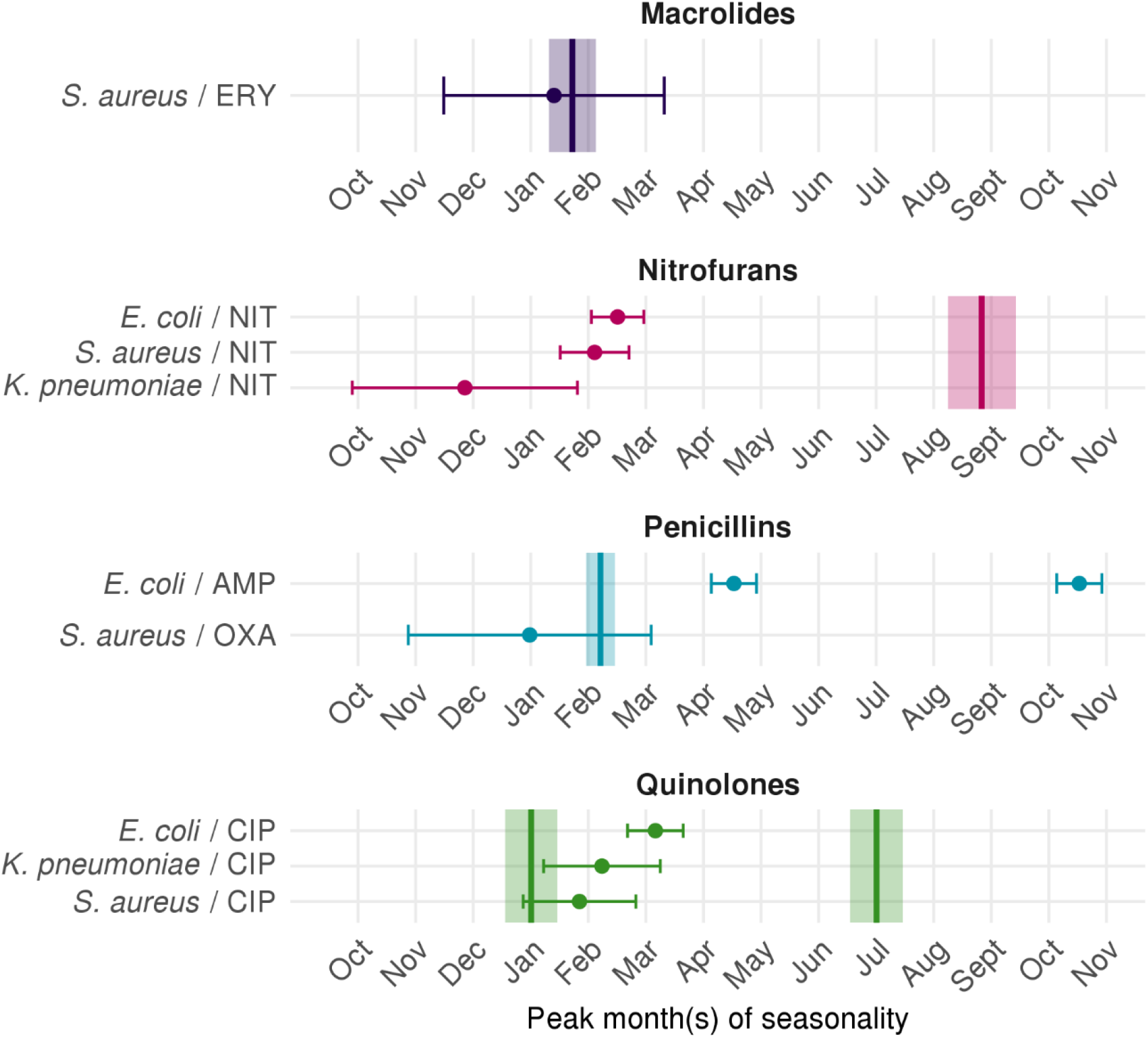
Phases of seasonality of use and resistance by species and antibiotic class. Points indicate peak month(s) of seasonal resistance estimated by the best-fitting sinusoidal model (comparing 6- and 12-month periods) for each species-antibiotic combination, and error bars indicate the 95% confidence intervals. Vertical lines indicate the peak month(s) of seasonal use estimated by the best-fitting sinusoidal model (comparing 6- and 12-month periods) for each antibiotic class, and the shaded regions indicate the 95% confidence intervals. Included are species-antibiotic combinations for which the amplitude was statistically significant (FDR < 0.05). AMC, Amoxicillin-Clavulanate; AMP, Ampicillin; CIP, Ciprofloxacin; ERY, Erythromycin; NIT, Nitrofurantoin; OXA, Oxacillin.

Since the antibiotic use dataset was restricted to outpatient prescribing for people under 65 years of age, we repeated the resistance analysis on data subset to isolates from outpatients under 65 years old, representing 53%, 31%, and 47% of the *E. coli, K. pneumoniae*, and *S. aureus* isolates, respectively. Resistance to ampicillin and nitrofurantoin in *E. coli* and nitrofurantoin in *S. aureus* remained seasonal with statistically significant seasonal amplitudes (FDR < 0.05) and showed the same periods and phases of seasonality as in the analyses with the full dataset including all isolates (**S1 Table**). Resistance to ciprofloxacin in *E. coli* also remained seasonal with a statistically significant amplitude in both the 12- and 6-month period model; however, a 6-month period model performed marginally better than the 12-month model (Akaike information criterion (AIC) difference, 0.2). In contrast, resistance to all antibiotics in *K. pneumoniae* and ciprofloxacin, erythromycin, and oxacillin resistance in *S. aureus* no longer met our criterion for seasonality (amplitude FDR < 0.05) after restricting our analysis to outpatients under 65 years old. For ciprofloxacin resistance with a 12-month period model and erythromycin resistance with a 6-month period model in *S. aureus*, the amplitude *p* values were < 0.05, but did not remain significant after multiple testing correction.

To further explore whether the observed seasonality of resistance could be attributable to seasonally varied sampling of patient demographics and sites of infection, we repeated the resistance analysis on the full dataset with all isolates after including covariates in our model to adjust for patient age and sex (Eqn. 3 in Materials and Methods) and patient age, sex, and site of infection (Eqn. 4 in Materials and Methods). Resistance remained seasonal for all 9 species-antibiotic combinations after adjusting for patient age and sex, with statistically significant amplitudes of seasonality (FDR < 0.05), though the magnitude of the estimated amplitudes decreased by 0-32% compared to the unadjusted model (**Table 1**). After adjusting for site of infection in addition to age and sex, resistance to ciprofloxacin, erythromycin, and oxacillin in *S. aureus* no longer met our criterion for seasonality (amplitude FDR > 0.05) and the estimated amplitudes decreased by 29-62% compared to the unadjusted model, while resistance in *E. coli* and *K. pneumoniae* remained seasonal (amplitude FDR < 0.05) with a 0-14% decrease in amplitude compared to the unadjusted model (**Table 1**). The amplitude *p* values were < 0.05 for ciprofloxacin and oxacillin resistance in *S. aureus* after adjusting for age, sex, and site of infection, but did not remain significant after multiple testing correction. In this model (Eqn. 4 in Materials and Methods), the coefficient for sex was significantly positive (FDR < 0.05; where a positive *β*_*s*_ indicates that being male is associated with higher MICs) across all antibiotics in *E. coli* (median, 0.30; range, 0.053 to 0.54) and *K. pneumoniae* (median, 0.25; range, 0.18 to 0.40) and largely non-significant in *S. aureus* (median, -2.3e-3; range, -0.079 to 0.020) (**S2 Table**). The coefficient for age was significantly positive (FDR < 0.05; where a positive *β*_*a*_ indicates that older ages are associated with higher MICs) in *E. coli* (median, 1.9e-3; range, 7.6e-4 to 0.013) and significantly in negative in all antibiotics except ciprofloxacin in *K. pneumoniae* (median, -1.0e-3; range, -2.7e-3 to 8.0e-4) (**S2 Table**). In *S. aureus*, the coefficient for age was significantly positive for ciprofloxacin, erythromycin, and oxacillin resistance, significantly negative for penicillin resistance, and non-significant for nitrofurantoin and tetracycline resistance (median for all antibiotics in *S. aureus*, 2.3e-3; range, -9.4e-4 to 0.016). Finally, at least one of the coefficients for site of infection was significant (FDR < 0.05) in all 15 species-antibiotic combinations, indicating that the site of infection was also an important determinant of MIC (**S2 Table**).

**Table 1.** Comparison of estimated amplitudes of seasonality across three sinusoidal models for resistance.

| Species | Abx | Period (months) | Amplitude (95% CI) |  |  |
| --- | --- | --- | --- | --- | --- |
|  |  |  | Model A (unadjusted) | Model B (adjusted for age and sex) | Model C (adjusted for age, sex, and site of infection) |
| *E. coli* | AMC | 6 | 0.01 (-5.1e-04, 0.021) | 0.011 (4.3e-04, 0.022) | 0.011 (6.3e-04, 0.022) |
| *E. coli* | AMP | 6 | 0.034 (0.019, 0.049) * | 0.034 (0.019, 0.048) * | 0.034 (0.019, 0.048) * |
| *E. coli* | CIP | 12 | 0.051 (0.031, 0.072) * | 0.045 (0.026, 0.065) * | 0.044 (0.025, 0.064) * |
| *E. coli* | NIT | 12 | 0.028 (0.02, 0.037) * | 0.028 (0.02, 0.036) * | 0.028 (0.02, 0.036) * |
| *E. coli* | TET | 6 | 0.013 (-3.6e-03, 0.029) | 0.013 (-3.4e-03, 0.029) | 0.013 (-3.3e-03, 0.029) |
| *K. pneumoniae* | AMC | 12 | 0.034 (1.2e-03, 0.067) | 0.034 (1.3e-03, 0.066) | 0.032 (-2.1e-04, 0.065) |
| *K. pneumoniae* | CIP | 12 | 0.053 (0.023, 0.083) * | 0.05 (0.02, 0.081) * | 0.048 (0.018, 0.078) * |
| *K. pneumoniae* | NIT | 12 | 0.035 (9.6e-03, 0.061) * | 0.034 (7.9e-03, 0.06) * | 0.034 (7.6e-03, 0.06) * |
| *K. pneumoniae* | TET | 6 | 0.021 (-9.8e-03, 0.051) | 0.019 (-0.011, 0.049) | 0.019 (-0.012, 0.049) |
| *S. aureus* | CIP | 12 | 0.063 (0.034, 0.093) * | 0.043 (0.016, 0.07) * | 0.024 (5.0e-04, 0.048) |
| *S. aureus* | ERY | 12 | 0.048 (0.012, 0.083) * | 0.042 (7.7e-03, 0.077) * | 0.032 (-1.0e-03, 0.065) |
| *S. aureus* | NIT | 12 | 0.033 (0.022, 0.043) * | 0.034 (0.023, 0.044) * | 0.038 (0.027, 0.048) * |
| *S. aureus* | OXA | 12 | 0.031 (8.8e-03, 0.054) * | 0.027 (5.5e-03, 0.049) * | 0.023 (8.2e-04, 0.044) |
| *S. aureus* | PEN | 6 | 0.01 (-6.6e-03, 0.027) | 9.5e-03 (-7.3e-03, 0.026) | 9.7e-03 (-7.1e-03, 0.027) |
| *S. aureus* | TET | 12 | 0.013 (-7.2e-03, 0.033) | 0.012 (-7.3e-03, 0.032) | 0.013 (-6.9e-03, 0.033) |

Model A does not adjust for the patient demographics or site of infection, Model B adjusts for patient age and sex, and Model C adjusts for patient age, sex, and site of infection. In parentheses are the 95% confidence intervals on the amplitude estimates. Asterisks indicate that the amplitude is significant after Benjamini-Hochberg multiple testing correction (false discovery rate < 0.05). AMC, Amoxicillin-Clavulanate; AMP, Ampicillin; CIP, Ciprofloxacin; ERY, Erythromycin; NIT, Nitrofurantoin; OXA, Oxacillin; PEN, Pencillin; TET, Tetracycline.

### Seasonal resistance is positively correlated with use of winter-peaking antibiotic classes

Spearman correlation coefficients between use-resistance antibiotic pairs varied widely across antibiotics, species, and lag times, ranging from -0.91 to 0.92 (**Fig 5, S4 Fig**). The number of statistically significant correlations between use-resistance pairs was maximized when the lag was 0 months in *S. aureus* and *K. pneumoniae* and 1 month in *E. coli*. Resistance across multiple antibiotics was most positively correlated with use of winter-peaking classes, penicillins and macrolides (median Spearman’s ρ across all lags, 0.45; interquartile range (IQR), 0.058 to 0.76). Resistance to most antibiotics also showed a negative correlation with use of summer-peaking classes, tetracyclines and nitrofurans (median Spearman’s ρ across all lags, -0.35; IQR, -0.63 to -0.068). Finally, resistance was not significantly correlated with use of quinolones, which peaked twice a year, for almost all antibiotics and species (median Spearman’s ρ across all lags, -0.21; IQR, -0.32 to -0.051).

**Fig 5.**
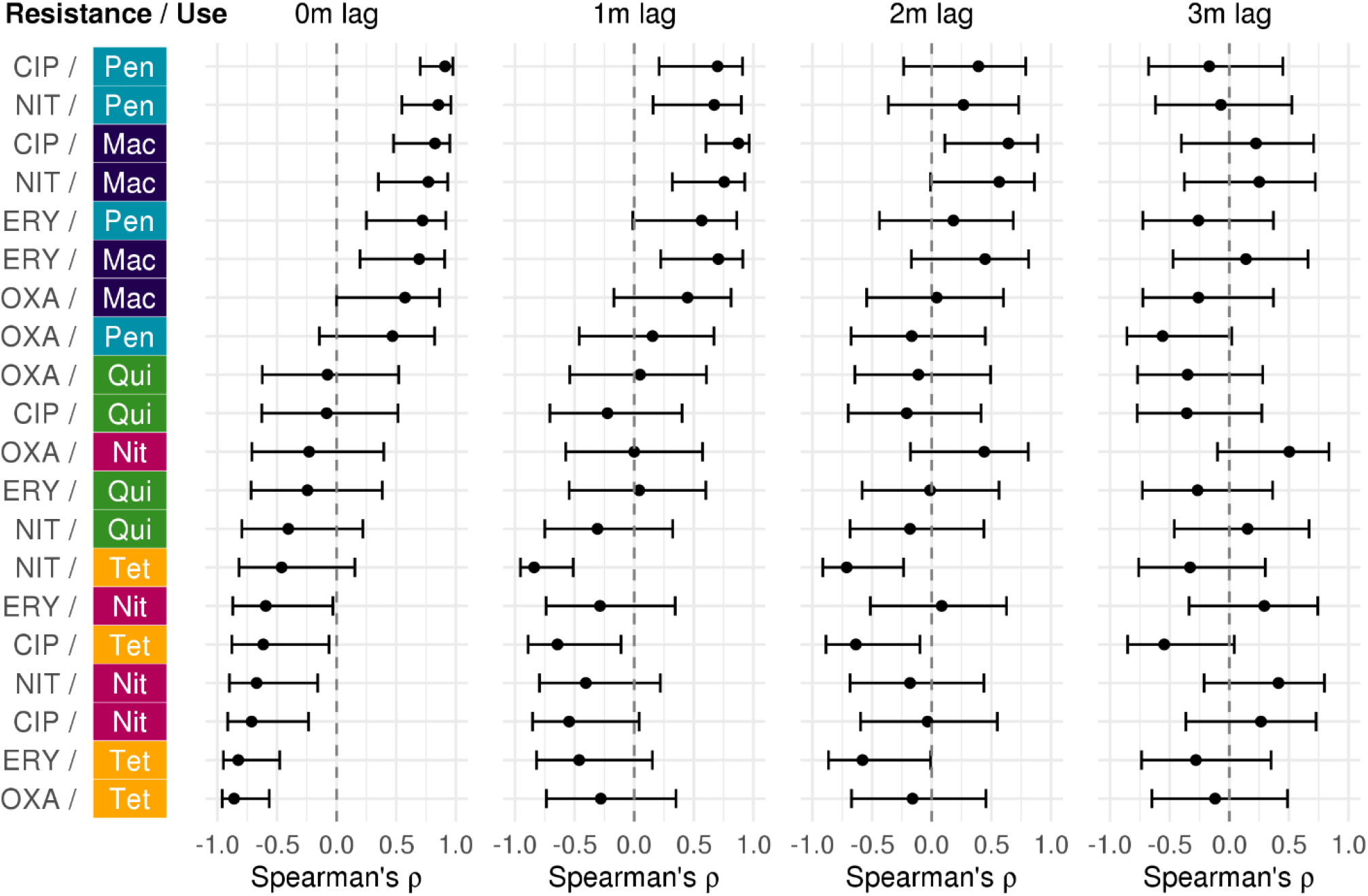
Spearman correlations between seasonal use and resistance with 0-3 months lag in *S. aureus*. Spearman’s rank correlation coefficients were calculated between the monthly mean seasonal deviate in resistance (in log_2_(MIC)) and the monthly mean seasonal deviate in use (in average daily claims per 10,000 people) with 0, 1, 2, or 3 months lag between use and resistance, for each pairwise combination of antibiotics and classes. Error bars indicate the 95% confidence intervals. Colors indicate the use antibiotic class. Mac, Macrolides; Nit, Nitrofurantoin; Pen, Penicillins; Qui, Quinolones; Tet, Tetracyclines. CIP, Ciprofloxacin; ERY, Erythromycin; NIT, Nitrofurantoin; OXA, Oxacillin.

## Discussion

Under a model in which antibiotic use drives resistance, seasonal variation in antibiotic consumption is expected to be associated with variation in population-level resistance that is in phase with or lagged up to a quarter period behind use [10]. However, we found that resistance to all antibiotics, including those with summer and biannual peaks in use, best correlated temporally with use of winter-peaking antibiotics—penicillins and macrolides—at a 0-to-1-month lag.

The observed patterns of use and resistance for penicillins and macrolides were mostly consistent with previous findings [6,7,9] and with model predictions [10]. Use of penicillins and macrolides peaked in the winter, likely due to increased wintertime prescribing for respiratory infections [12]. In *S. aureus*, resistance to oxacillin and erythromycin peaked in the winter and was most correlated with penicillins and macrolides use with no lag. This was consistent with a study that compared seasonal macrolide use and resistance in methicillin-resistant *S. aureus* (MRSA) [7] and findings in other species-antibiotic combinations that have shown winter peaks in use and resistance with little to no lag [6–8]. In *E. coli*, the first peak in ampicillin resistance occurred in the spring, lagging penicillins use by about 2.3 months, but also showed a second peak in resistance 6 months later, in the absence of a second peak in penicillins use. However, we note that a 12-month period model for ampicillin resistance in *E. coli* also met our criterion for seasonality and showed a single winter peak, which is more consistent with previous findings in *E. coli* [7,8].

Antibiotic classes with different seasonal patterns of use, such as nitrofurans and quinolones, showed seasonal patterns of resistance inconsistent with model predictions and not readily explained based on their patterns of use. Nitrofurans, which are almost exclusively used to treat urinary tract infections (UTIs) [16], showed summer peaks in use during the same season as peak UTI incidence [17–20]. However, resistance to nitrofurantoin in all 3 species peaked in the winter and lagged use by 3.2 to 5.7 months. This was consistent with a report that nitrofurantoin resistance lagged use by 3-6 months in *E. coli* and *K. pneumoniae* urinary tract isolates [11]. Quinolones, which are used to treat both respiratory infections and UTIs [21], showed both winter and summer peaks in use, but only a single winter/spring peak in ciprofloxacin resistance in all 3 species.

Several species-antibiotic combinations did not show seasonality in resistance. In each case, this may be due to a lack of association between use and resistance, a lack of strong seasonal variation in use of some antibiotic classes, or other factors, such as a signal too small to be identified or dampened by the combination of inpatient and outpatient samples in our dataset. The lack of seasonality in *S. aureus* resistance to penicillin may be explained by the high prevalence of penicillin resistance in *S. aureus* [22] (83% in this dataset), which may limit the observable effect of increased wintertime use of penicillins. For resistance to amoxicillin-clavulanate in *E. coli* and *K. pneumoniae*, the estimated amplitude of seasonality missed the 5% FDR cutoff for statistical significance, but additional data may narrow the confidence intervals around the estimated amplitude. The absence of observed seasonality in tetracycline resistance in all 3 species is consistent with a previous study that found no significant correlation between tetracycline use and resistance in *E. coli* [7] and may be explained by the lack of strong seasonal variation in tetracycline use (**Fig 1**).

Our finding that resistance to all antibiotics most correlated with use of winter-peaking antibiotic classes suggests that the simple model in which use of a given antibiotic independently selects for resistance is insufficient to explain the full seasonal use-resistance landscape. Below, we discuss four factors that may contribute to this result, while acknowledging that there may be additional unknown seasonally varying determinants of resistance.

First, the lack of association between seasonal use and resistance in some antibiotics may be an artifact of comparing between use and resistance in overlapping but not identical populations. Data availability limited us to comparing between antibiotic use in outpatients under age 65 and resistance measured in inpatients and outpatients at two hospitals with patient populations that skew towards older ages. Comparisons between population-level community use and hospital resistance have frequently been utilized in previous ecological [23–27] and seasonal [6–8,11] studies, due to common difficulties in both obtaining within-hospital use data and tracking community infections that do not result in a healthcare visit. The volume of antibiotic use in the community is much greater than in the hospital [28] and thus can have a strong impact on resistance in both settings [29,30]. This impact appears to vary across species-antibiotic combinations and by demographics. For quinolones, we observed biannual peaks in use and winter peaks in resistance in all three pathogens; it may be that the populations receiving quinolones in the summer and winter are not sampled equally in our resistance dataset. Including only outpatients under 65 years old in our resistance analysis resulted in the loss of statistically significant seasonality in *S. aureus* and *K. pneumoniae* for some antibiotics. While this may be attributable to reduction in signal (this subset represents only 30-50% of the total isolates), this result could also suggest that the observed seasonal trends in resistance for some species-antibiotic combinations are disproportionately driven by older populations with infections associated with hospitalization. In contrast, the seasonal patterns of resistance among outpatients under 65 largely remained the same across antibiotics in *E. coli* and for nitrofurantoin in *S. aureus*. Thus, the disparity in community and hospital use is unlikely to fully explain the lack of association in seasonal patterns of use and resistance.

Second, the observed seasonal peaks in resistance could be driven by seasonal variation in the incidence of infection with a given species, although no specific mechanism for this association has been proposed to our knowledge. In our dataset, isolate counts peaked in the summer for all three species considered (**S5 Fig**), while most resistance peaked in the winter. Therefore, for seasonal variations in incidence to drive seasonality in resistance, there would need to be an inverse association between incidence and resistance; however, since there is no proposed mechanism for this association, we did not explore it statistically.

In contrast, a third mechanism by which resistance could vary seasonally is that certain patient demographics, or certain sites of infection, are associated with resistance and themselves vary seasonally. Rates of resistance have been shown to vary by age, sex, and site of infection [31– 33], and, the incidence of infections from these groups have been shown to vary by season in our dataset (**S5 Fig**) and others [20,34]. If these factors fully explained seasonal variation in resistance, incorporating them as covariates in our seasonal regression (assuming the association was correctly specified) should have accounted for the seasonal signal and led to near-zero estimates for the sinusoidal component of the regression. However, resistance remained seasonal with amplitudes decreasing by 0-32%, but remaining statistically significant, in all 9 species-antibiotic combinations after accounting for age and sex (**Table 1**). After also accounting for site of infection, the seasonal amplitudes of resistance in 6 of those 9 combinations remained significant, decreasing by 0-14% compared to the unadjusted model. However, for the 3 antibiotics in *S. aureus* where resistance was no longer seasonal, the amplitudes decreased substantially by 29-62% compared to the unadjusted model. Therefore, seasonally varied sampling of isolates from different demographic groups and sites of infection likely contributes to but does not fully explain the observed seasonality in resistance.

Fourth, the winter peaks in resistance to antibiotics with different seasonal peaks in use could be explained by co-selection, where use of one antibiotic can indirectly select for resistance to a second antibiotic in bacteria that are co-resistant to both antibiotics [35]. Co-resistance between penicillins/macrolides and other antibiotics is common across many bacterial species, including those in our study [36]. Therefore, the winter peaks in resistance to other antibiotics may be driven by co-selection by winter-peaking use of penicillins and macrolides. We might expect that selection by use of pencillins and macrolides dominates over selection by other antibiotics because they are prescribed at substantially higher rates and show greater seasonal variations in use [12]. Antibiotics with higher rates of use showed stronger correlations between seasonal use and resistance [7]. In addition, use of macrolides and penicillins have been shown to be more strongly correlated with resistance than less frequently prescribed antibiotics [23].

There were several limitations to this study. First, we measured antibiotic use in the population by the number of claims per capita, rather than daily doses, making the assumption that the average dose and duration do not vary greatly within the short timescales in which we are measuring seasonal variations in use, and that there were not major selective differences between the effects of one prescription for different members of an antibiotic class. Second, we were unable to link antibiotic prescriptions to specific pathogens, and thus we could not assess the extent to which antibiotic resistance in a given species is attributable to antibiotic use for the treatment of infections caused by that species. However, given that bystander selection has been predicted to account for over 80% of the total antibiotic selection experienced by *S. aureus, E. coli*, and *K. pneumoniae* [3], the analysis we performed comparing total antibiotic use to resistance in each of these species may in fact yield more relevant interpretations of the use-resistance relationship [37]. Third, the antibiotic use and resistance datasets that were available to us for each species and antibiotic often spanned overlapping but different year ranges. Therefore, we aggregated monthly use and resistance data across years to perform our correlation analyses. We accounted for variability in use and resistance between years by adjusting for annual trends in use and resistance in our model.

In conclusion, this work contributes to describing the complexity of the antibiotic use-resistance relationship. Our finding that resistance to all antibiotics peaked in the winter/spring, regardless of patterns of use, is consistent with studies from a range of geographic scales and regions, including South Carolina [8], the USA overall [7,9], Israel [6], and the Netherlands [11]. This suggests a general phenomenon in which selection or co-selection by those antibiotics with high-volume, winter-peaking use and/or other ecological factors results in wintertime peaks of resistance for all antibiotics and indicates that the simplest model of antibiotic use independently driving resistance to the same antibiotic is inadequate. We show that additional factors, such as seasonal variations in the contribution of isolates from different demographic groups, may also contribute to the observed winter peaks in resistance. This study lays the groundwork for future work to further identify and describe the factors that shape the use-resistance landscape across diverse pathogens and antibiotics, with important implications for informing and monitoring the outcome of efforts to reduce antibiotic resistance.

## Materials and Methods

### Antibiotic use data

Outpatient antibiotic use data was obtained from the Massachusetts All Payer Claims Database [38], which covers >94% of outpatient prescriptions claims for Massachusetts residents under the age of 65 [39]. Rates of use for each antibiotic class were measured as the average daily number of antibiotic claims per 10,000 people during each calendar month from January 2011 to May 2015. This data was subset to include only individuals residing in ‘Boston City’ census tracks, as defined by the US Census Bureau [40], to capture the antibiotic use patterns in the communities served by the hospitals in our resistance dataset. We aggregated antibiotic use data by class according to the World Health Organization’s Anatomical Therapeutic Chemical Classification System [41] (**S3 Table**). We included 5 antibiotic classes in our analysis, which together make up 74% of the total outpatient antibiotic claims in Boston: penicillins, macrolides, quinolones, tetracyclines, and nitrofurans. Given that bystander selection likely accounts for most of the antibiotic selection experienced by *S. aureus, E. coli*, and *K. pneumoniae* [3], we included use data for all antibiotics within each class, regardless of the target pathogen for which they were prescribed.

### Antibiotic resistance data

Clinical microbiology data was obtained for *S. aureus, E. coli*, and *K. pneumoniae* isolates collected at two tertiary care hospitals in Boston, MA: Brigham and Women’s Hospital (BWH) and Massachusetts General Hospital (MGH), from 2007-2019 and 2007-2016, respectively. Included in this analysis were all non-surveillance isolates from inpatients and outpatients of all demographics, collected from the 5 most common sites of infection across the 3 species: blood, skin and soft tissue, abscess/fluid, respiratory tract, and urinary tract (**S4 Table, S5 Fig**). Isolates of the same species that were collected from the same patient within 2 weeks were assumed to represent a single infection and thus treated as a single isolate. Our final dataset comprised of 47,374 *S. aureus*, 130,407 *E. coli*, and 27,178 *K. pneumoniae* isolates.

Antibiotic susceptibility testing was performed on each isolate either by automated broth microdilution (Vitek 2, Biomerieux, Marcy l’Etoile, France) or by gradient diffusion. Resulting minimum inhibitory concentration (MIC) values were log_2_-transformed. When MICs were reported with an inequality sign, we used only the numerical value in our quantitative analyses. Due to variations in hospital testing guidelines across the years, we excluded tests on isolates that did not report an MIC value, either because a different test method was used (e.g., disk diameter) or due to missing data. We excluded years/months from our analysis for each species-antibiotic combination in each hospital where MIC values were reported for fewer than 80% of isolates or only a subset of isolate types (e.g., only testing nitrofurantoin resistance in urinary tract isolates). **S5 Table** lists date ranges and percent resistance in each hospital, calculated as the percentage of non-susceptible isolates out of the total number of isolates from each hospital with a reported MIC value, for each species-antibiotic combination included in our analysis. In S5 Table, we determined antibiotic susceptibility by applying the Clinical & Laboratory Standards Institute 2017 breakpoints [42] to the reported MIC values of each isolate; the determined susceptibility was then adjusted based on β-lactamase screen and cefoxitin screen results if available for penicillin and oxacillin, respectively. For all other analyses other than in S5 Table, we use the log_2_-transformed MIC as the unit of resistance. This study was approved by the Mass General Brigham Institutional Review Board (Protocol number: 2016P001671).

### Statistical methods

We quantified the extent of seasonality in antibiotic use and resistance by fitting the use and MIC data to a pair of mathematical models, based on a previously described method [9,10]. Both models consist of (a) a sinusoidal component to describe seasonal deviations from average year-round use and MICs and (b) a linear component to adjust for secular trends, such as declines in use and resistance across years [22,39]. This model makes no assumptions about the underlying mechanism of resistance and is generalizable to any species and antibiotic [9,10]. As in the Olesen et al. model [9], we chose to substitute MIC for the original outcome (proportion resistance) of the Blanquart et al. model [10] to allow for detection of seasonal variations in the quantitative level of resistance, as measured by MIC, even if that variation occurs without crossing a defined breakpoint for antibiotic susceptibility.

To describe the seasonality of use, monthly claims data for each antibiotic class were fit to

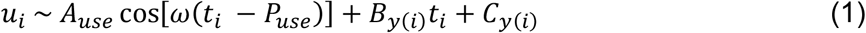

where *u*_*i*_ is the mean daily reported claims per 10,000 people during calendar month *t*_*i*_, *A*_*use*_ is the amplitude of use seasonality, *ω* is the frequency of seasonality where 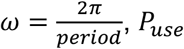 is the phase of use seasonality, and *B*_*y*_(_*i*_) and *C*_*y*_(_*i*_) are the within-year slope and intercept terms. To describe the seasonality of resistance, MICs for each isolate were fit to

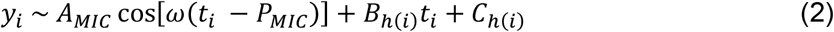

where *y*_*i*_ is the log_2_-transformed MIC and *t*_*i*_ is the calendar month of collection of the *i*^*th*^ isolate, *A*_*MIC*_ is the amplitude of resistance seasonality, *ω* is the frequency of seasonality where 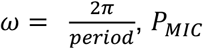 is the phase of resistance seasonality, and *B*_*h*_(_*i*_) and *C*_*h*_(_*i*_) are the within hospital/year slope and intercept terms.

We further fit the MIC data to two additional models to account for the effect of seasonally varied sampling of patient demographics and sites of infection on the observed seasonality of resistance. To describe the seasonality of resistance while adjusting for patient demographics, age and sex, we fit the MIC data to

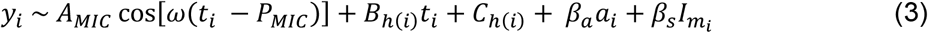

where *β*_*a*_ and *β*_*s*_ are the coefficients for age and sex, respectively, *a*_*i*_ is the age (in years) of the patient from which the *i*^*th*^ isolate is collected, and 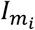 is an indicator variable for whether the patient is male. To describe the seasonality of resistance while adjusting for patient demographics and site of infection, we fit the MIC data to

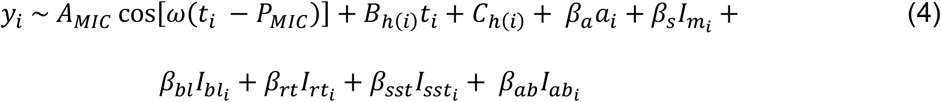

where *β*_*bl*_, *β*_*rt*_, *β*_*sst*_, and *β*_*ab*_ are the coefficients for blood, respiratory tract, skin and soft tissue, and abscess/fluid, respectively, and 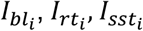, and 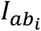 are the indicator variables for whether the *i*^*th*^ isolate was collected from the blood, respiratory tract, skin and soft tissue, or abscess/fluid, respectively.

The amplitude, phase, slope, intercept, and demographic coefficient terms in each model were estimated by non-linear regression, using the *nls* function in R (version 3.6.2) [43]. We examined periods of both 12 and 6 months to account for annual or biannual cycles in use and resistance. We justified using these fixed periods by performing a wavelet analysis, using the *WaveletComp* package [44] in R, on the raw antibiotic use data to show that the dominant periods of variations in use across the included years are at 12 and 6 months (**S6 Fig**). To determine whether to use a 12- or 6-month period for each species-antibiotic combination, we performed model comparisons using the Akaike information criterion (AIC) and used the period that resulted in the lower AIC (**S6 Table, S7 Table**). We determined that there was seasonality in use or resistance if the amplitude was statistically significant after accounting for multiple comparisons by applying the Benjamini-Hochberg correction with a 5% false discovery rate (FDR).

We quantified the association between the observed seasonal patterns of use and resistance using Spearman’s rank correlations. To eliminate the impact of annual trends, we calculated correlations between the average monthly seasonal deviates in use and resistance, aggregated across years, rather than the raw use and MIC data. We define a ‘seasonal deviate’ as the deviation in use or MIC at a given time of year from the year-round average, which we estimated by the linear component of the models. Seasonal deviates in use for each year and month were calculated as

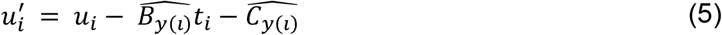

where 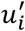 is the seasonal deviate of the mean reported daily claims per 10,000 people during calendar month *t*_*i*_, *u*_*i*_ is the mean reported daily claims per 10,000 people during calendar month *t*_*i*_, and 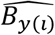 and 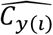 are the within-year slope and intercept terms estimated from the model fit (Eqn. 1) for the corresponding year. For resistance, we calculate the seasonal deviate for each isolate as

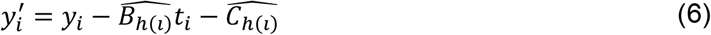

where 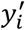 is the seasonal deviate of the log_2_-transformed MIC of the *i*^*th*^ isolate, *y*_*i*_ is the log_2_-transformed MIC of the *i*^*th*^ isolate, *t*_*i*_ is the calendar month of collection of the *i*^*th*^ isolate, and 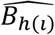 and 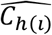 are the hospital/year slope and intercept estimated from the model fit (Eqn. 2) for the hospital and year that the *i*^*th*^ isolate was collected in.

Since the working model for the use-resistance relationship predicts that seasonal fluctuations in resistance can lag use by up to 3 months [10], we calculated Spearman correlations between use and resistance seasonal deviates with no lag and lags of 1, 2, and 3 months. In addition, because we observed some seasonal patterns of resistance that better aligned with use of non-cognate antibiotic classes, we calculated use-resistance correlations between each pairwise combination of target antibiotics and use classes. We only included use-resistance pairs in this analysis for which both use and resistance met our criterion for seasonality.

All analyses were performed in R version 3.6.2 [43]. Data and code are available at: https://github.com/gradlab/use-resistance-seasonality

## Data Availability

Code and data is available at: https://github.com/gradlab/use-resistance-seasonality

## Acknowledgements

The authors thank R. Monina Klevens for assistance in acquiring the antibiotic prescribing data and Enterprise Research Infrastructure & Services at Partners HealthCare for their computational resources and support.

---

**S1 Fig.**
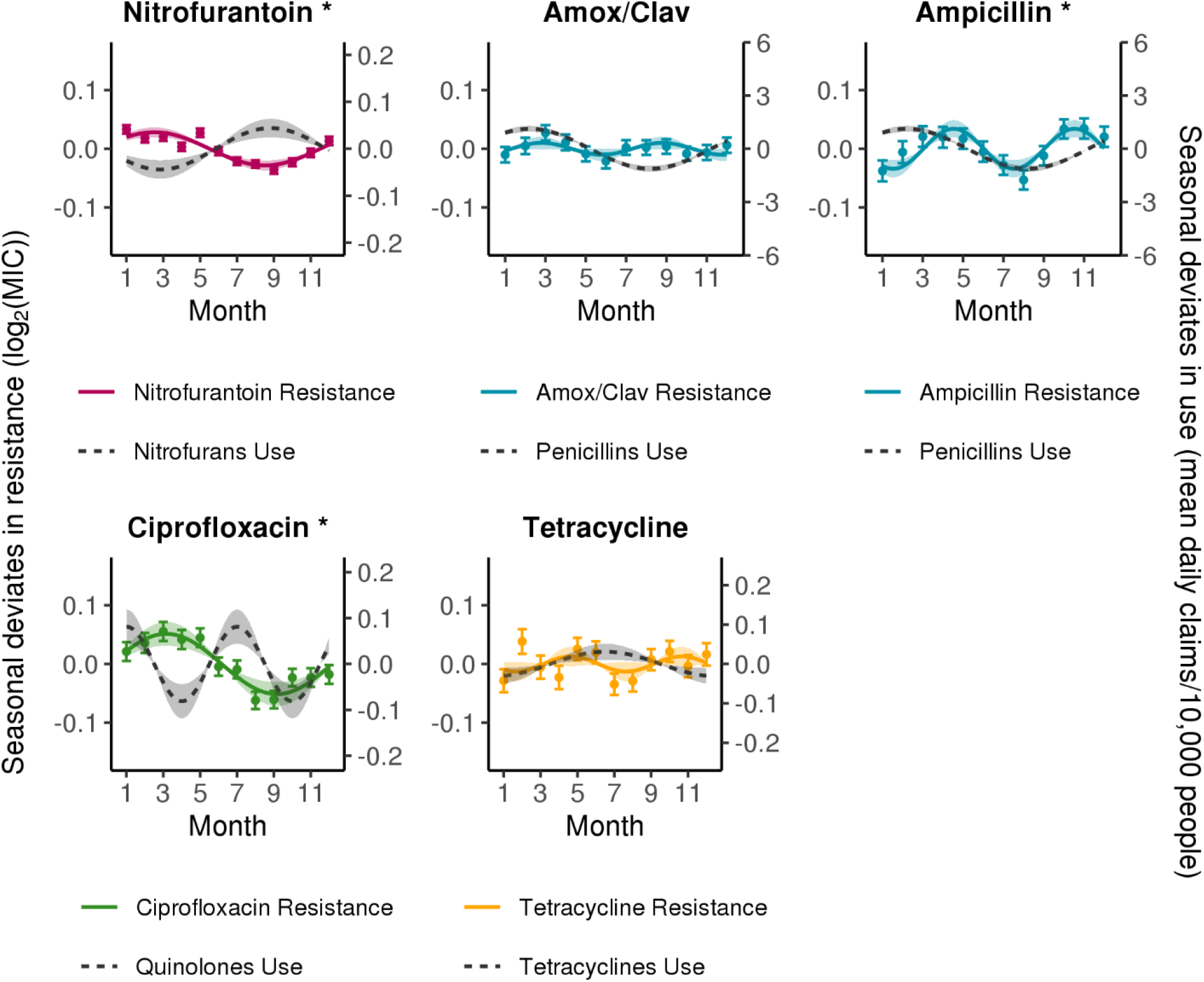
Seasonality of antibiotic use and resistance by class in *Escherichia coli*. Solid lines indicate point estimates of the amplitude and phase from the best-fitting sinusoidal model of resistance (comparing 6- and 12-month periods) to each antibiotic, colored by class. Dashed grey lines indicate point estimates of the amplitude and phase from sinusoidal models of use of the corresponding antibiotic class. Shaded regions indicate the 95% confidence intervals for the amplitude. Points indicate the monthly mean seasonal deviates in resistance and error bars indicate the standard error of the mean. Asterisks indicate the amplitude of seasonality in resistance is statistically significant (FDR < 0.05). Amox/Clav, Amoxicillin-Clavulanate.

**S2 Fig.**
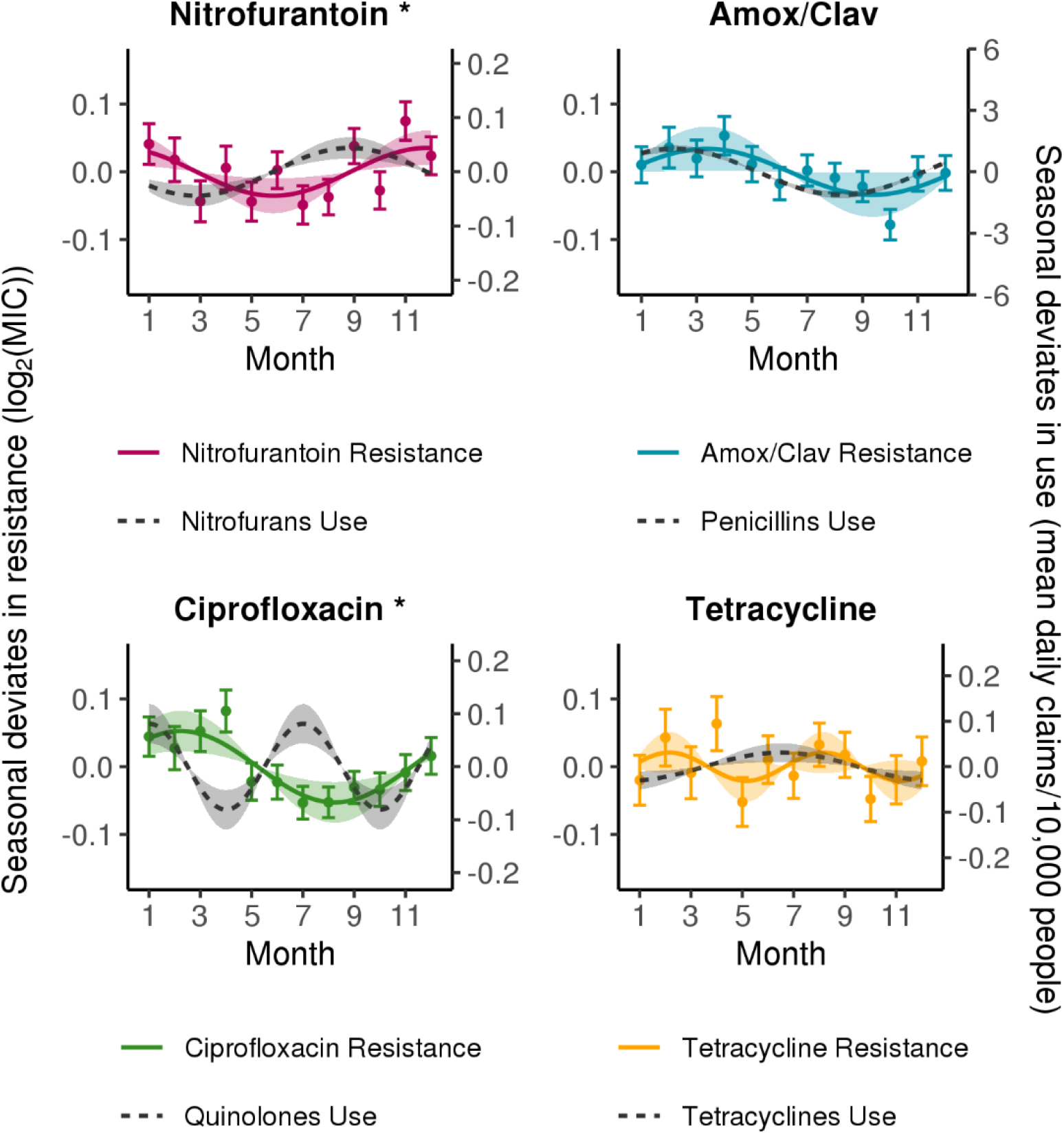
Seasonality of antibiotic use and resistance by class in *Klebsiella pneumoniae*. Solid lines indicate point estimates of the amplitude and phase from the best-fitting sinusoidal model (comparing 6- and 12-month periods) of resistance to each antibiotic, colored by class. Dashed grey lines indicate point estimates of the amplitude and phase from sinusoidal models of use of the corresponding antibiotic class. Shaded regions indicate the 95% confidence intervals for the amplitude. Points indicate the monthly mean seasonal deviates in resistance and error bars indicate the standard error of the mean. Asterisks indicate the amplitude of seasonality in resistance is statistically significant (FDR < 0.05). Amox/Clav, Amoxicillin-Clavulanate.

**S3 Fig.**
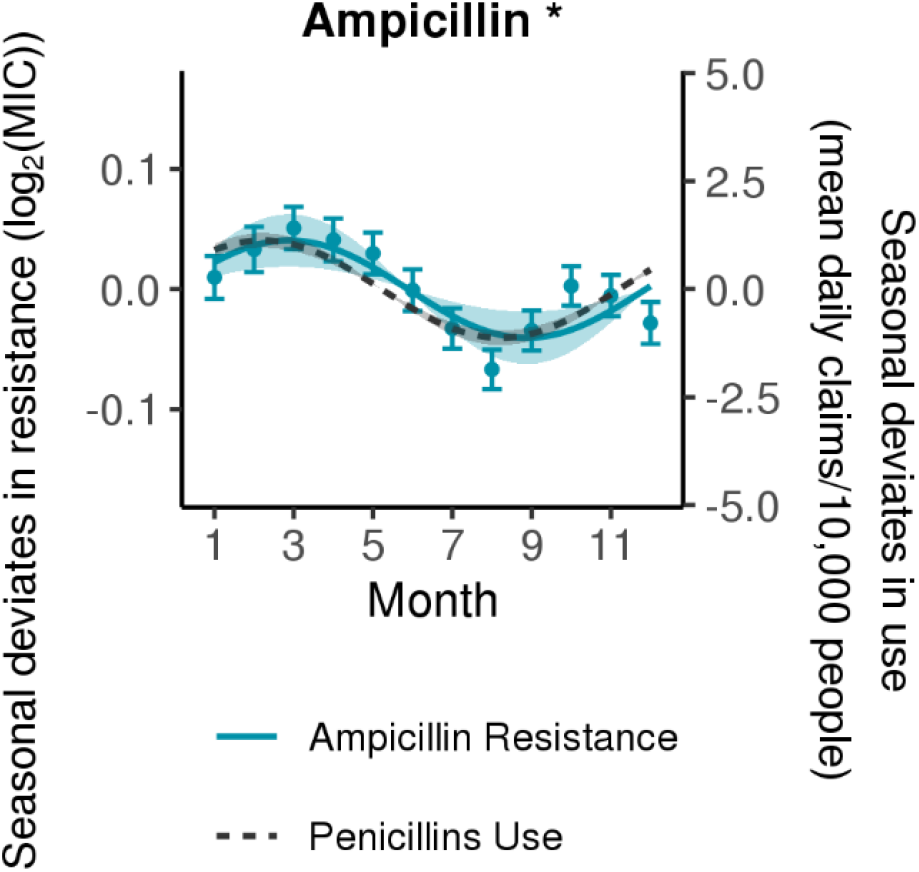
Seasonality of penicillins use and ampicillin resistance in *Escherichia coli* with a 12-month period model. Solid line indicates point estimates of the amplitude and phase from a 12-month period sinusoidal model of resistance to ampicillin in *E. coli*. Dashed grey line indicates point estimates of the amplitude and phase from a 12-month period sinusoidal model of use of penicillin class antibiotics. Shaded regions indicate the 95% confidence intervals for the amplitude. Points indicate the monthly mean seasonal deviates in resistance and error bars indicate the standard error of the mean. Asterisk indicates the amplitude of seasonality in resistance is statistically significant (FDR < 0.05).

**S4 Fig.**
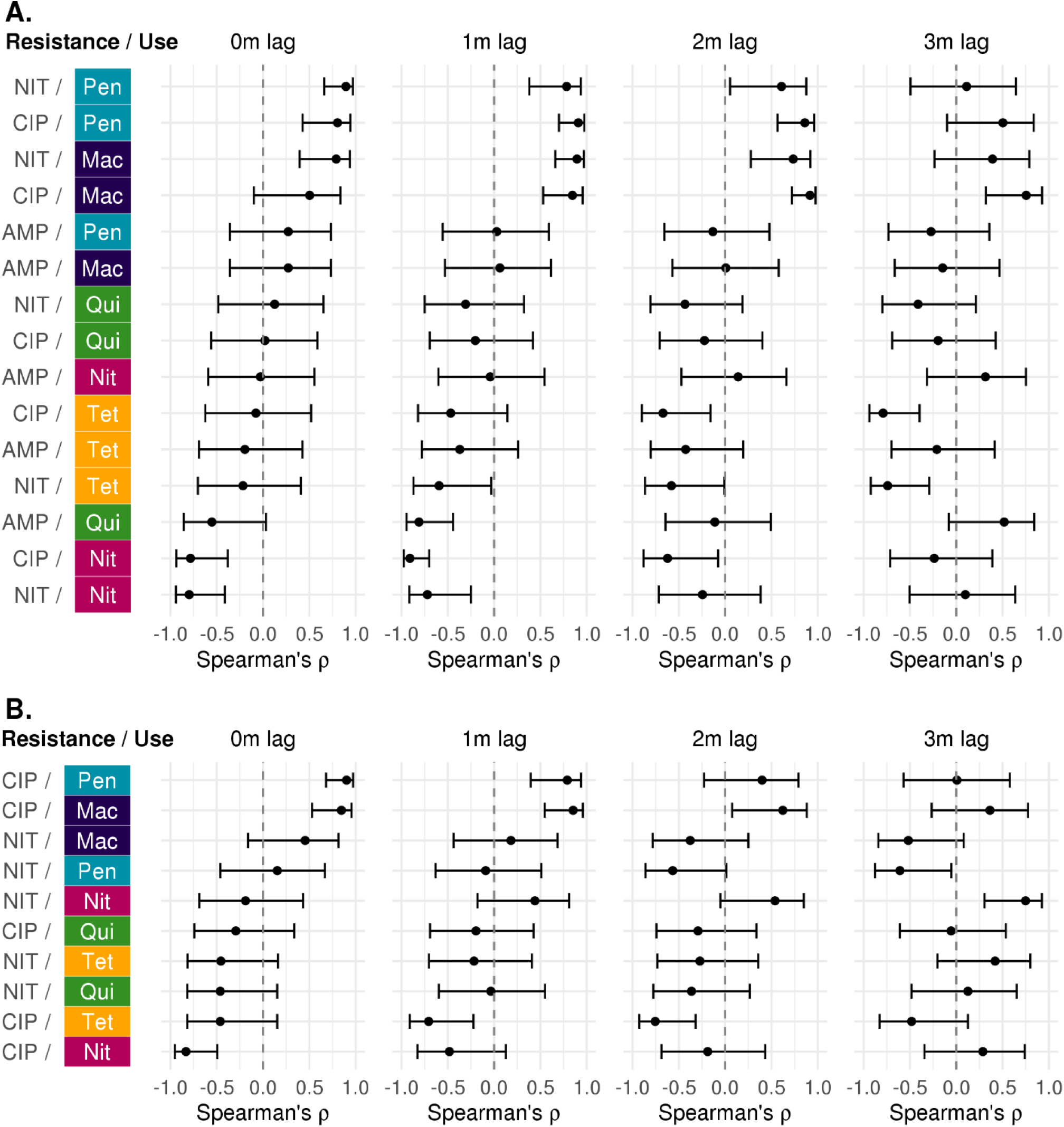
Spearman correlations between seasonal use and resistance with 0-3 months lag in (A) *E. coli* and (B) *K. pneumoniae*. Spearman’s rank correlation coefficients were calculated between the monthly mean seasonal deviate in resistance (in log_2_(MIC)) and the monthly mean seasonal deviate in use (in average daily claims per 10,000 people) with 0, 1, 2, or 3 months lag between use and resistance, for each pairwise combination of antibiotics and classes. Error bars indicate the 95% confidence intervals. Colors indicate the use antibiotic class. Mac, Macrolides; Nit, Nitrofurantoin; Pen, Penicillins; Qui, Quinolones; Tet, Tetracyclines. AMP, Ampicillin; CIP, Ciprofloxacin; NIT, Nitrofurantoin.

**S5 Fig.**
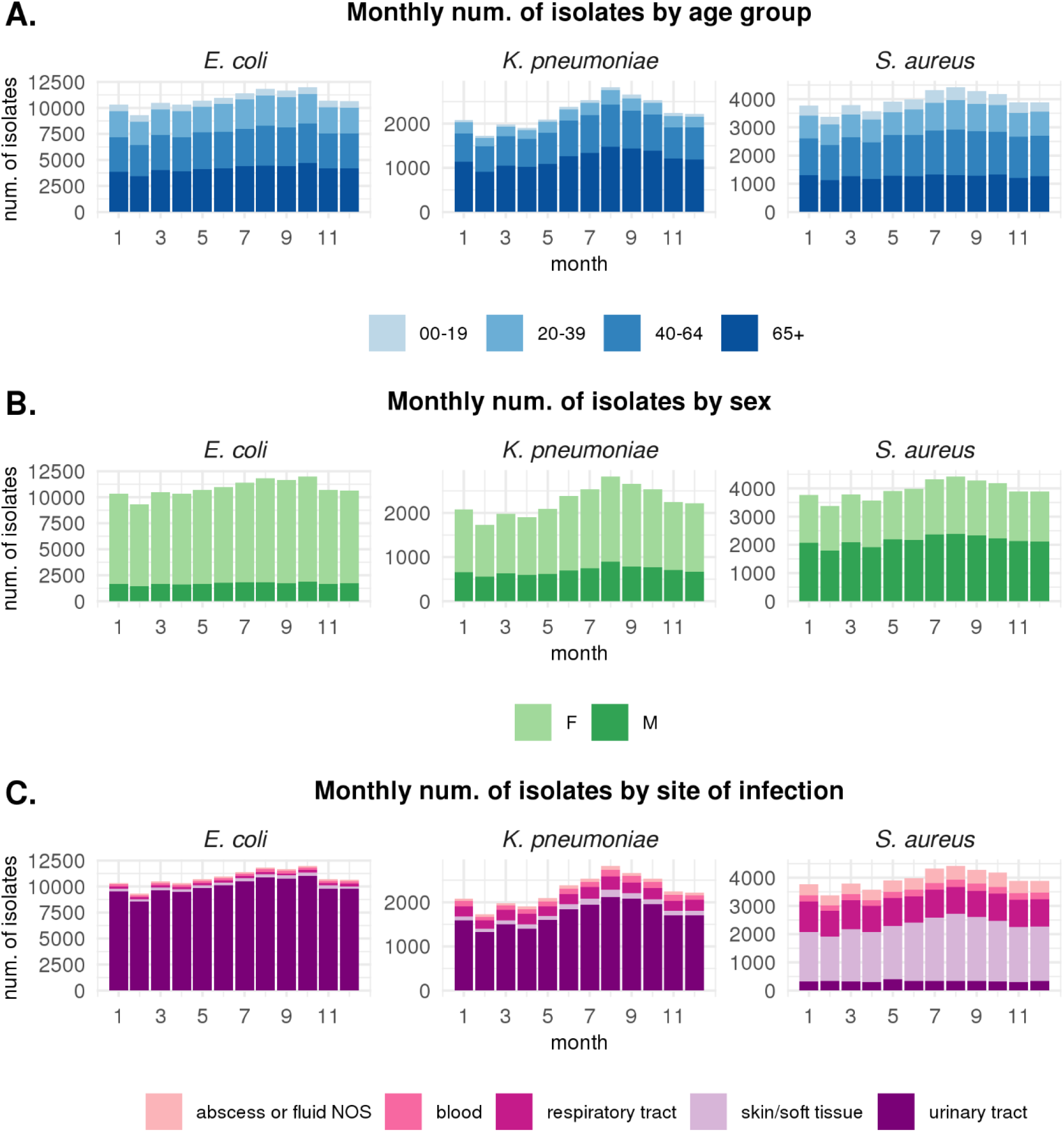
Seasonal incidence of infection by demographic group or site of infection. Bars show the total number of isolates by month included in the resistance dataset for each species, colored by (A) age group, (B) sex, and (C) site of infection. NOS, not otherwise specified.

**S6 Fig.**
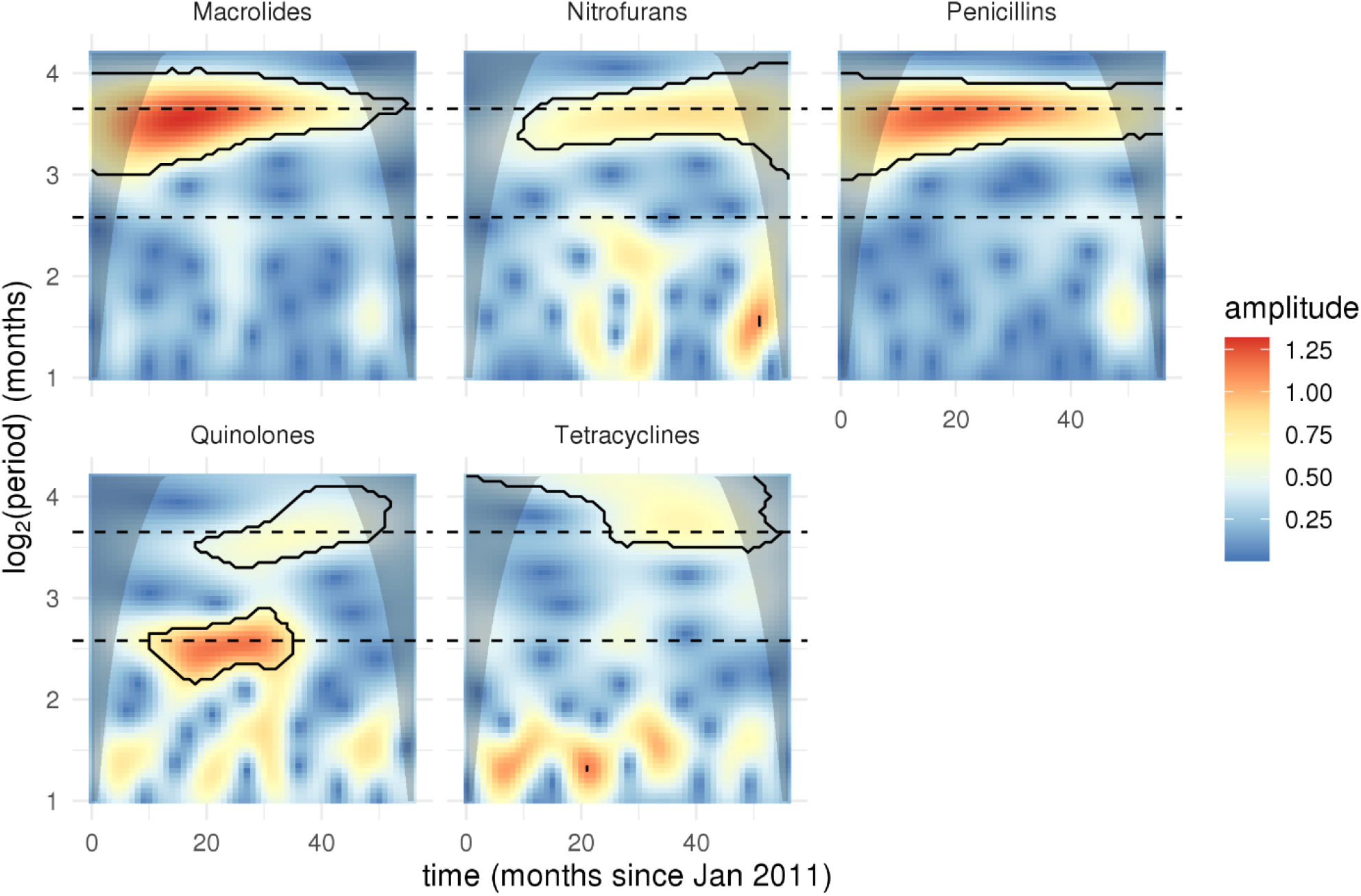
Wavelet analysis of antibiotic use by class. Dotted lines show 12-month (upper line) and 6-month (lower line) periods. Solid lines indicate regions where the amplitude *p* value is less than 0.05. Shaded areas indicate the ‘cone of influence’ where edge effects are important.

**S1 Table.** Amplitudes and phases of seasonality of resistance in outpatients under 65 years old.

| Species | Antibiotic | Period | Amplitude (95% CI) | Phase (95% CI) |
| --- | --- | --- | --- | --- |
| *E. coli* | AMC | 6 months | 0.013 (-1.7e-03, 0.027) | 2.7 (1.6, 3.7) |
| *E. coli* | AMP | 6 months | 0.039 (0.019, 0.06) * | 4.3 (3.8, 4.8) |
| *E. coli* | CIP | 6 months | 0.022 (5.9e-03, 0.037) * | 4.6 (3.9, 5.2) |
| *E. coli* | NIT | 12 months | 0.033 (0.022, 0.044) * | 2.6 (2.1, 3.1) |
| *E. coli* | TET | 6 months | 0.013 (-7.9e-03, 0.034) | 3.6 (2, 5.3) |
| *K. pneumoniae* | AMC | 12 months | 0.028 (-0.027, 0.082) | 4 (1.6, 6.4) |
| *K. pneumoniae* | CIP | 6 months | 0.023 (-0.014, 0.059) | 4.8 (3.4, 6.2) |
| *K. pneumoniae* | NIT | 6 months | 0.027 (-0.013, 0.067) | 5.5 (4.1, 7) |
| *K. pneumoniae* | TET | 12 months | 0.062 (-0.012, 0.14) | 8.3 (6.2, 10) |
| *S. aureus* | CIP | 12 months | 0.043 (2.2e-03, 0.085) | 2.2 (0.46, 3.9) |
| *S. aureus* | ERY | 6 months | 0.056 (8.9e-03, 0.1) | 2.6 (1.8, 3.4) |
| *S. aureus* | NIT | 12 months | 0.041 (0.025, 0.058) * | 2.4 (1.7, 3.1) |
| *S. aureus* | OXA | 12 months | 0.03 (-0.017, 0.078) | 11 (8.2, 13) |
| *S. aureus* | PEN | 12 months | 0.031 (-5.7e-03, 0.069) | 9.1 (7.5, 11) |
| *S. aureus* | TET | 12 months | 0.019 (-8.6e-03, 0.047) | 11 (8.4, 13) |

Amplitudes and phases were estimated from the best-fitting sinusoidal model of resistance (comparing 6- and 12-month periods) for each species-antibiotic combination, using data that was subset to include only outpatients under age 65. Asterisks indicate that the amplitude is significant after Benjamini-Hochberg multiple testing correction (false discovery rate < 0.05). AMC, Amoxicillin-Clavulanate; AMP, Ampicillin; CIP, Ciprofloxacin; ERY, Erythromycin; NIT, Nitrofurantoin; OXA, Oxacillin; PEN, Pencillin; TET, Tetracycline.

**S2 Table.**
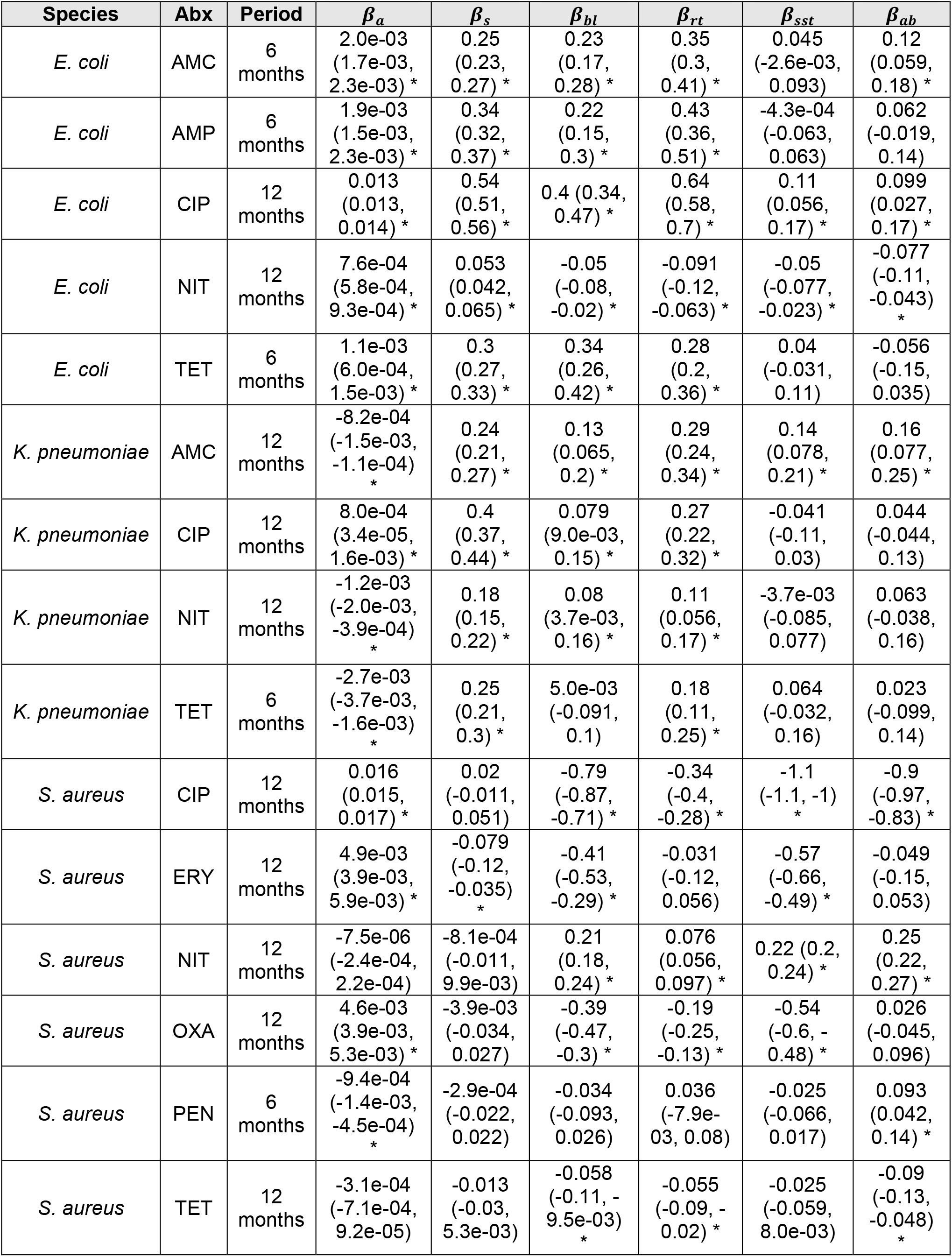
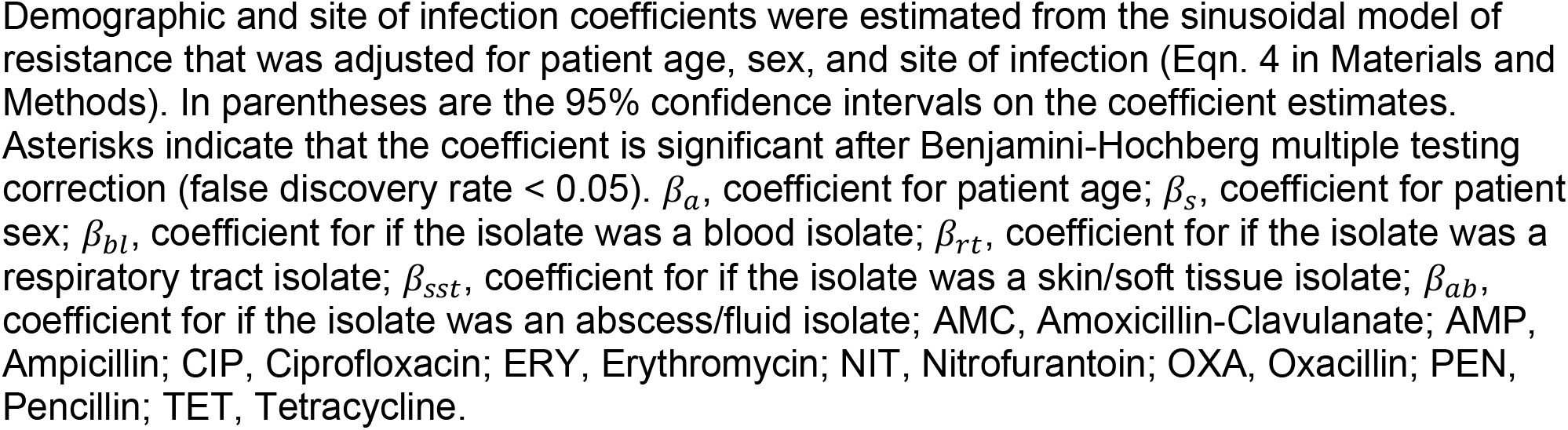
Age, sex, and site of infection coefficients for adjusted sinusoidal model of seasonal resistance.

**S3 Table.**
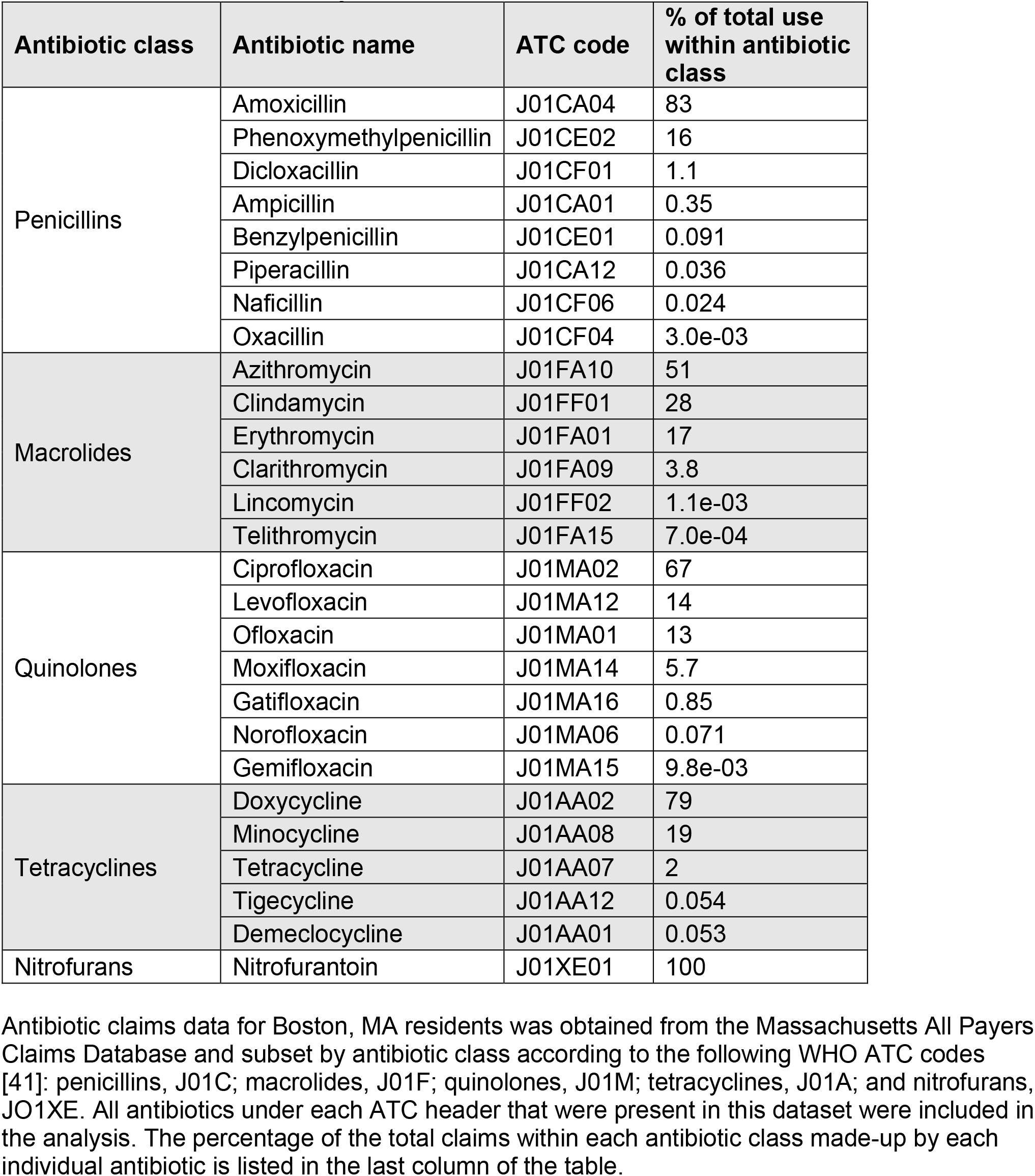
Percent of claims by individual antibiotics within each class.

**S4 Table.**
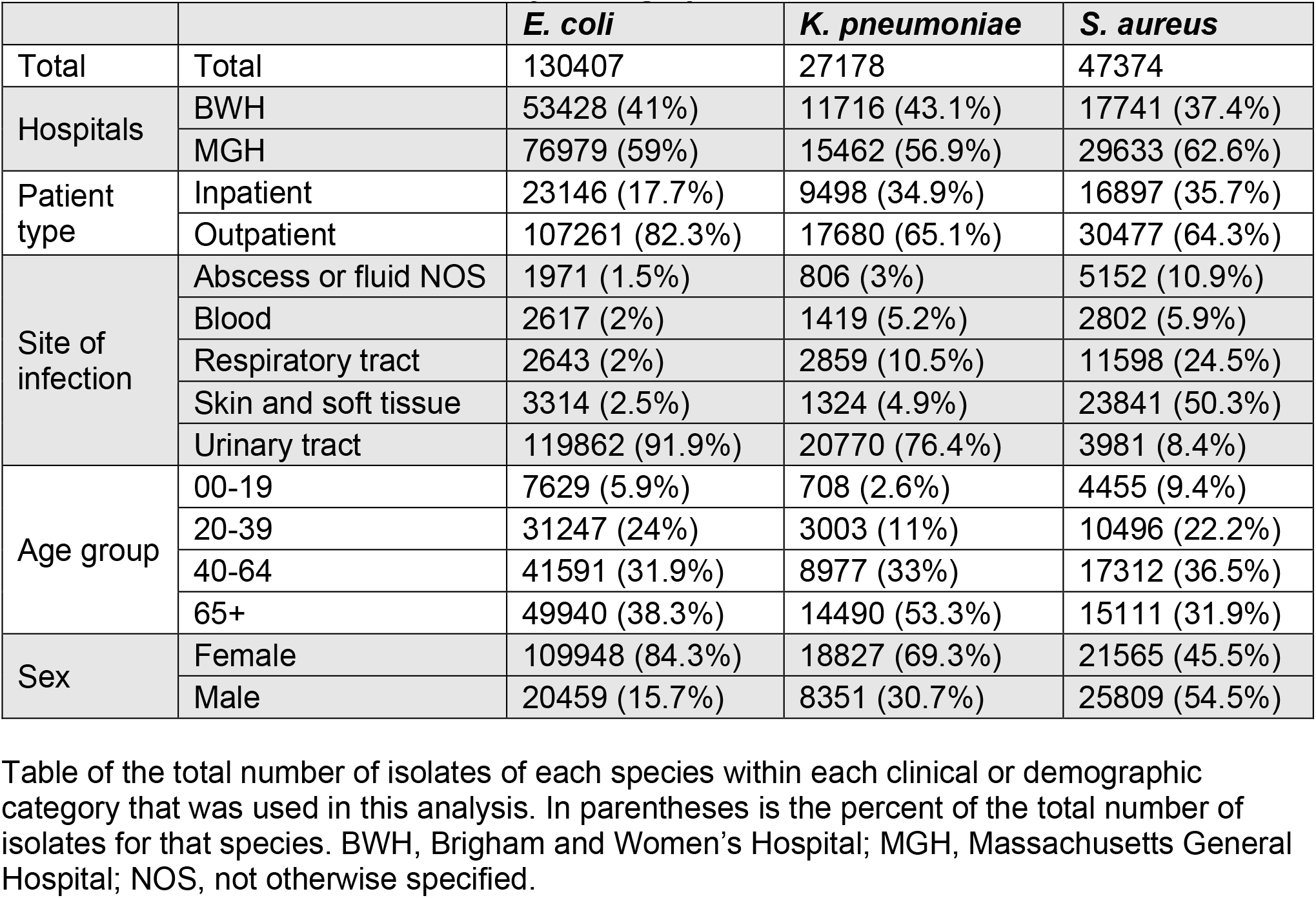
Total number of isolates by demographics.

|  |  | *E. coli* | *K. pneumoniae* | *S. aureus* |
| --- | --- | --- | --- | --- |
| Total | Total | 130407 | 27178 | 47374 |
| Hospitals | BWH | 53428 (41%) | 11716 (43.1%) | 17741 (37.4%) |
|  | MGH | 76979 (59%) | 15462 (56.9%) | 29633 (62.6%) |
| Patient type | Inpatient | 23146 (17.7%) | 9498 (34.9%) | 16897 (35.7%) |
|  | Outpatient | 107261 (82.3%) | 17680 (65.1%) | 30477 (64.3%) |
| Site of infection | Abscess or fluid NOS | 1971 (1.5%) | 806 (3%) | 5152 (10.9%) |
|  | Blood | 2617 (2%) | 1419 (5.2%) | 2802 (5.9%) |
|  | Respiratory tract | 2643 (2%) | 2859 (10.5%) | 11598 (24.5%) |
|  | Skin and soft tissue | 3314 (2.5%) | 1324 (4.9%) | 23841 (50.3%) |
|  | Urinary tract | 119862 (91.9%) | 20770 (76.4%) | 3981 (8.4%) |
| Age group | 00-19 | 7629 (5.9%) | 708 (2.6%) | 4455 (9.4%) |
|  | 20-39 | 31247 (24%) | 3003 (11%) | 10496 (22.2%) |
|  | 40-64 | 41591 (31.9%) | 8977 (33%) | 17312 (36.5%) |
|  | 65+ | 49940 (38.3%) | 14490 (53.3%) | 15111 (31.9%) |
| Sex | Female | 109948 (84.3%) | 18827 (69.3%) | 21565 (45.5%) |
|  | Male | 20459 (15.7%) | 8351 (30.7%) | 25809 (54.5%) |

Table of the total number of isolates of each species within each clinical or demographic category that was used in this analysis. In parentheses is the percent of the total number of isolates for that species. BWH, Brigham and Women's Hospital; MGH, Massachusetts General Hospital; NOS, not otherwise specified.

**S5 Table.** Antibiotics included in analysis and percent resistance by hospital.

| Species | Antibiotic<br>(Antibiotic class) | Dates<br>included at<br>BWH | Dates<br>included at<br>MGH | Percent<br>resistance<br>at BWH | Percent<br>resistance<br>at MGH |
| --- | --- | --- | --- | --- | --- |
| *E. coli* | Amoxicillin/<br>Clavulanate<br>(Penicillins) | May 2013 –<br>Dec 2019 | Jan 2007 –<br>Dec 2016 | 17.8% | 17.2% |
|  | Ampicillin<br>(Penicillins) | Jan 2007 –<br>Dec 2019 | Jan 2007 –<br>Dec 2016 | 47.8% | 47.0% |
|  | Ciprofloxacin<br>(Quinolones) | Jan 2007 –<br>Dec 2019 | Jan 2007 –<br>Dec 2016 | 22.6% | 21.1% |
|  | Nitrofurantoin<br>(Nitrofurans) | Jan 2007 –<br>Jun 2013 | Jan 2007 –<br>Dec 2016 | 6.0% | 5.1% |
|  | Tetracycline<br>(Tetracyclines) | May 2013 –<br>Dec 2019 | Jan 2007 –<br>Dec 2016 | 29.1% | 29.3% |
| *K. pneumoniae* | Amoxicillin/<br>Clavulanate<br>(Penicillins) | May 2013 –<br>Dec 2019 | Jan 2007 –<br>Dec 2016 | 10.2% | 8.2% |
|  | Ciprofloxacin<br>(Quinolones) | Jan 2007 –<br>Dec 2019 | Jan 2007 –<br>Dec 2016 | 11.8% | 11.1% |
|  | Nitrofurantoin<br>(Nitrofurans) | Jan 2007 –<br>Jun 2013 | Jan 2007 –<br>Dec 2016 | 69.4% | 68.0% |
|  | Tetracycline<br>(Tetracyclines) | May 2013 –<br>Dec 2019 | Jan 2007 –<br>Dec 2016 | 21.3% | 18.3% |
| *S. aureus* | Ciprofloxacin<br>(Quinolones) | May 2010 –<br>Dec 2019 | Jan 2009 –<br>Dec 2016 | 29.6% | 30.7% |
|  | Erythromycin<br>(Macrolides) | May 2010 –<br>Dec 2019 | Jan 2009 –<br>Dec 2016 | 51.2% | 54.9% |
|  | Nitrofurantoin<br>(Nitrofurans) | May 2010 –<br>Jun 2013 | Jan 2009 –<br>Dec 2016 | 1.1% | 1.0% |
|  | Oxacillin<br>(Penicillins) | May 2010 –<br>Dec 2019 | Jan 2009 –<br>Dec 2016 | 33.4% | 35.9% |
|  | Penicillin<br>(Penicillins) | May 2010 –<br>Dec 2019 | Jan 2009 –<br>Apr 2015 | 82.4% | 84.3% |
|  | Tetracycline<br>(Tetracyclines) | May 2010 –<br>Dec 2019 | Jan 2009 –<br>Dec 2016 | 6.6% | 6.0% |

Percent resistance was calculated as the percentage of non-susceptible isolates out of the total number of isolates from each hospital with a reported minimum inhibitory concentration for that antibiotic. BWH, Brigham and Women's Hospital; MGH, Massachusetts General Hospital.

**S6 Table.** Comparison of the Akaike information criterion (AIC) values between 6- and 12-month period models for antibiotic use.

| Antibiotic class | Antibiotic Use Regression Model |  |
| --- | --- | --- |
|  | 6-month period | 12-month period |
| Macrolides | 104.4 (+67.5) | 36.9 (+0) |
| Nitrofurans | -201.0 (+12.1) | -213.1 (+0) |
| Penicillins | 128.7 (+86.6) | 42.0 (+0) |
| Quinolones | -89.5 (+0) | -79.6 (+9.9) |
| Tetracyclines | -155.1 (+6.1) | -161.2 (+0) |

In parentheses is the difference in AIC from the model with the lower AIC.

**S7 Table.**
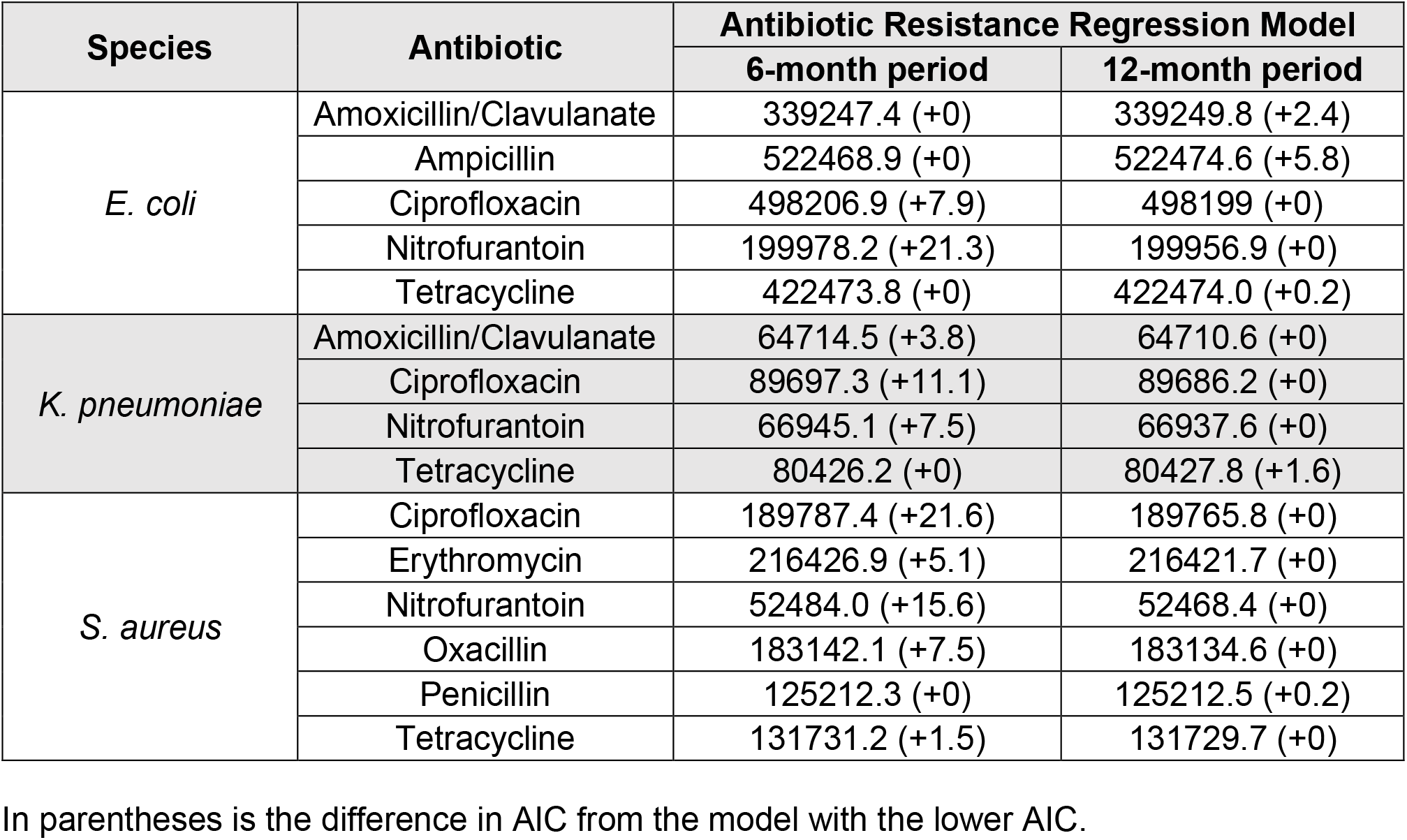
Comparison of the AIC values between 6- and 12-month period models for antibiotic resistance.

## References

1. U.S. Centers for Disease Control and Prevention. Antibiotic resistance threats in the United States. 2019. Available from: https://www.cdc.gov/drugresistance/pdf/threats-report/2019-ar-threats-report-508.pdf

2. Jit M, Ng DHL, Luangasanatip N, Sandmann F, Atkins KE, Robotham JV, et al. Quantifying the economic cost of antibiotic resistance and the impact of related interventions: Rapid methodological review, conceptual framework and recommendations for future studies. BMC Med. 2020;18(1):38.

3. Tedijanto C, Olesen SW, Grad YH, Lipsitch M. Estimating the proportion of bystander selection for antibiotic resistance among potentially pathogenic bacterial flora. Proc Natl Acad Sci USA. 2018;115(51):E11988–95.

4. Hennessy TW, Petersen KM, Bruden D, Parkinson AJ, Hurlburt D, Getty M, et al. Changes in antibiotic-prescribing practices and carriage of penicillin-resistant Streptococcus pneumoniae: A controlled intervention trial in rural Alaska. Clin Infect Dis. 2002;34(12):1543–50.

5. Enne VI, Livermore DM, Stephens P, Hall LMC. Persistence of sulphonamide resistance in Escherichia coli in the UK despite national prescribing restriction. Lancet. 2001;357(9265):1325–28.

6. Dagan R, Barkai G, Givon-Lavi N, Sharf AZ, Vardy D, Cohen T, et al. Seasonality of Antibiotic-Resistant Streptococcus pneumoniae That Causes Acute Otitis Media: A Clue for an Antibiotic-Restriction Policy? J Infect Dis. 2008;197(8):1094–102.

7. Sun L, Klein EY, Laxminarayan R. Seasonality and Temporal Correlation between Community Antibiotic Use and Resistance in the United States. Clin Infect Dis. 2012;55(5):687–94.

8. Ramsey EG, Royer J, Bookstaver PB, Justo JA, Kohn J, Albrecht H, et al. Seasonal variation in antimicrobial resistance rates of community-acquired Escherichia coli bloodstream isolates. Int J Antimicrob Agents. 2019;54(1):1–7.

9. Olesen SW, Torrone EA, Papp JR, Kirkcaldy RD, Lipsitch M, Grad YH. Azithromycin Susceptibility Among Neisseria gonorrhoeae Isolates and Seasonal Macrolide Use. J Infect Dis. 2019;219(4):619–23.

10. Blanquart F, Lehtinen S, Fraser C. An evolutionary model to predict the frequency of antibiotic resistance under seasonal antibiotic use, and an application to Streptococcus pneumoniae. Proc R Soc B Biol Sci. 2017;284:20170679.

11. Martínez EP, van Rosmalen J, Bustillos R, Natsch S, Mouton JW, Verbon A. Trends, seasonality and the association between outpatient antibiotic use and antimicrobial resistance among urinary bacteria in the Netherlands. J Antimicrob Chemother. 2020;75(8):2314–25.

12. Suda KJ, Hicks LA, Roberts RM, Hunkler RJ, Taylor TH. Trends and Seasonal Variation in Outpatient Antibiotic Prescription Rates in the United States, 2006 to 2010. Antimicrob Agents Chemother. 2014;58(5):2763–6.

13. Kluytmans J, Van Belkum A, Verbrugh H. Nasal Carriage of I: Epidemiology, Underlying Mechanisms, and Associated Risks. Clin Microbiol Rev. 1997;10(3):505–20.

14. Huttenhower C, Gevers D, Knight R, et al. Structure, function and diversity of the healthy human microbiome. Nature. 2012;486(7402):207–14.

15. Berchick ER, Barnett JC, Upton RD. Current population reports, P60-267 (RV), Health Insurance Coverage in the United States: 2018. Washington, DC: US Government Printing Office, 2019. Available at: https://www.census.gov/content/dam/Census/library/publications/2019/demo/p60-267.pdf

16. Guay DR. An Update on the Role of Nitrofurans in the Management of Urinary Tract Infections. Drugs. 2001;61(3):353–64.

17. Stamm WE, McKevitt M, Roberts PL, White NJ. Natural History of Recurrent Urinary Tract Infections in Women. Rev Infect Dis. 1991;13(1):77–84.

18. Anderson JE. Seasonality of symptomatic bacterial urinary infections in women. J Epidemiol Community Health. 983;37:286–90.

19. Czaja CA, Scholes D, Hooton TM, Stamm WE. Population-Based Epidemiologic Analysis of Acute Pyelonephritis. Clin Infect Dis. 2007;45(3):273–80.

20. Rosello A, Pouwels KB, Domenech De Cellès M, Van Kleef E, Hayward AC, Hopkins S, et al. Seasonality of urinary tract infections in the United Kingdom in different age groups: Longitudinal analysis of The Health Improvement Network (THIN). Epidemiol Infect 2018;146(1):37–45.

21. Hooper DC. Clinical applications of quinolones. Biochim Biophys Acta. 1998;1400(1):45–61.

22. Kanjilal S, Sater MRA, Thayer M, Lagoudas GK, Kim S, Blainey PC, et al. Trends in Antibiotic Susceptibility in Staphylococcus Aureus in Boston, Massachusetts, from 2000 to 2014. J Clin Microbiol. 2018;56(1):e01160–17.

23. Olesen SW, Barnett ML, Macfadden DR, Brownstein JS, Hernandez-Diaz S, Lipsitch M, et al. The distribution of antibiotic use and its association with antibiotic resistance. eLife. 2018;7:e39435.

24. Hicks LA, Chien Y-W, Taylor TH, Haber M, Klugman KP. Outpatient Antibiotic Prescribing and Nonsusceptible Streptococcus pneumoniae in the United States, 1996– 2003. Clin Infect Dis. 2011;53(7):631–9.

25. Bronzwaer SLAM, Cars O, Buchholz U, Molstad S, Goettsch W, Veldhuijzen IK, et al. The relationship between antimicrobial use and antimicrobial resistance in Europe. Emerg Infect Dis. 2002;8(3):278–82.

26. Bergman M, Huikko S, Pihlajamäki M, Laippala P, Palva E, Huovinen P, et al. Effect of macrolide consumption on erythromycin resistance in Streptococcus pyogenes in Finland in 1997-2001. Clin Infect Dis. 2004;38(9):1251–6.

27. MacDougall C, Powell JP, Johnson CK, Edmond MB, Polk RE. Hospital and community fluoroquinolone use and resistance in Staphylococcus aureus and Escherichia coli in 17 US hospitals. Clin Infect Dis. 20015;41(4):435–40.

28. ESPAUR Writing Committee. English Surveillance Programme for Antimicrobial Utilisation and Resistance (ESPAUR) Report 2020-2021. Public Health England. 2021. Available from: https://assets.publishing.service.gov.uk/government/uploads/system/uploads/attachment_data/file/1033851/espaur-report-2020-to-2021-16-Nov.pdf

29. Knight GM, Costelloe C, Deeny SR, Moore LSP, Hopkins S, Johnson AP, et al. Quantifying where human acquisition of antibiotic resistance occurs: A mathematical modelling study. BMC Med. 2018;16(1):137.

30. Macfadden DR, Fisman DN, Hanage WP, Lipsitch M. The Relative Impact of Community and Hospital Antibiotic Use on the Selection of Extended-spectrum Beta-lactamase-producing Escherichia coli. Clin Infect Dis. 2019;69(1):182–8.

31. Mcgregor JC, Elman MR, Bearden DT, Smith DH. Sex-and age-specific trends in antibiotic resistance patterns of Escherichia coli urinary isolates from outpatients. BMC Fam Pract. 2013;14:25.

32. Garcia A, Delorme T, Nasr P. Patient age as a factor of antibiotic resistance in methicillin-resistant Staphylococcus aureus. J Med Microbiol. 2017;66(12):1782–9.

33. Lagacé-Wiens PRS, DeCorby MR, Baudry PJ, Hoban DJ, Karlowsky JA, Zhanel GG. Differences in antimicrobial susceptibility in Escherichia coli from Canadian intensive care units based on regional and demographic variables. Can J Infect Dis Med Microbiol. 2008;19(4):282–6.

34. Leekha S, Diekema DJ, Perencevich EN. Seasonality of staphylococcal infections. Clin Microbiol Infec.. 2012;18(10):927–33.

35. Pouwels KB, Muller-Pebody B, Smieszek T, Hopkins S, Robotham J V. Selection and co-selection of antibiotic resistances among Escherichia coli by antibiotic use in primary care: An ecological analysis. PLOS One. 2019;14(6):e0218134.

36. Chang H-H, Cohen T, Grad YH, Hanage WP, O’Brien TF, Lipsitch M. Origin and Proliferation of Multiple-Drug Resistance in Bacterial Pathogens. Microbiol Mol Biol Rev. 2015;79(1):101–16.

37. Schechner V, Temkin E, Harbarth S, Carmeli Y, Schwaber MJ. Epidemiological Interpretation of Studies Examining the Effect of Antibiotic Usage on Resistance. Clin Microbiol Rev. 2013;26(2):289.

38. Center for Health Information and Analysis. Massachusetts All Payer Claims Database. 2019. Available from: https://www.chiamass.gov/ma-apcd/

39. Klevens RM, Caten E, Olesen SW, DeMaria A, Troppy S, Grad YH. Outpatient Antibiotic Prescribing in Massachusetts, 2011–2015. Open Forum Infect Dis. 2019;6(5):ofz159.

40. US Census Bureau. American fact finder. 2020. Available from: https://data.census.gov/cedsci/

41. World Health Organization Collaborating Centre for Drug Statistics Methodology. Structure and Principles. 2020. Available from: https://www.whocc.no/atc/structure_and_principles/

42. Clinical and Laboratory Standards Institute. Performance Standards for Antimicrobial Susceptibility Testing. 27th ed. CLSI M100. Wayne, PA. 2017.

43. R Core Team. R: A language and environment for statistical computing. R Foundation for Statistical Computing, Vienna, Austria.

44. Roesch A, Schmidbauer H. WaveletComp. 2018. Available from: https://cran.r-project.org/web/packages/WaveletComp/WaveletComp.pdf

